# Evaluating the Efficacy of Miglustat and Ambroxol Combination Therapy in a Phase 1b/2 Virtual Drug Trial for Type 1 Tay-Sachs Disease Using aiHumanoid Simulations

**DOI:** 10.1101/2024.11.21.24317709

**Authors:** WR Danter

## Abstract

Type 1 Tay-Sachs Disease (TSD) is a rare and severe neurodegenerative disorder caused by HEXA Loss of Function (LOF) gene mutations, leading to the accumulation of lysosomal GM2 gangliosides and progressive neurological decline, with high lethality before age 10. Current treatments are primarily palliative, focusing on symptom management. This study investigates the therapeutic potential of Miglustat and Ambroxol, individually and combined, in slowing neurodegeneration and cognitive decline in Type 1 TSD, leveraging aiHumanoid virtual simulations. Miglustat, a substrate reduction therapy, and Ambroxol, a pharmacological chaperone, offer complementary mechanisms that may enhance lysosomal function and reduce ganglioside accumulation. In a virtual Phase 1b/2 trial, simulated cohorts of children received these treatments across four dose levels (10%, 20%, 33.3%, and 50% of maximum tolerated dose [MTD]), with outcomes assessed at intervals from birth to 10 years. Key metrics included cognitive and motor function, quality of life, and lysosomal enzyme activity. Results indicated that combination therapy significantly reduced neurodegenerative symptoms and improved cognitive and motor outcomes, particularly at intermediate dose levels. Findings suggest that Miglustat and Ambroxol may provide a beneficial intervention strategy, warranting further clinical evaluation. These results also provide important potential insights into dose optimization and therapeutic synergies, offering a basis for real-world trials in Type 1 Tay-Sachs Disease. The use of aiHumanoid simulations demonstrates a novel approach for drug testing in rare diseases, enabling detailed assessment of efficacy and safety profiles across developmental stages. The trial’s findings are based on virtual simulations rather than traditional clinical trials.

## Introduction

Tay-Sachs Disease (TSD) is an autosomal recessive neurodegenerative disorder caused by loss of function (LOF) mutations in the HEXA gene, which result in a deficiency of β-hexosaminidase A, an enzyme involved in breaking down GM2 gangliosides. (lipids that can accumulate in nerve cells). This enzyme deficiency leads to GM2 ganglioside buildup within neurons, causing progressive neurological damage, particularly in Type 1 TSD, the most severe form of the disease [1,8]. Type 1 TSD is marked by rapid cognitive and motor deterioration, typically resulting in death before the age of 10, with no current cure and limited treatment options [2]. Due to the rare nature of the disorder combined with its high lethality, aiHumanoid simulations of TSD offer unique potential for gaining early insights into therapeutic strategies for rare diseases like TSD [6,7]. Traditionally, clinical trials for rare diseases often face challenges such as limited patient populations and recruitment delays that lead to cost, time delays and eventual failure. Virtual drug trials address these limitations by providing a scalable and ethically sound platform for evaluating therapeutic efficacy, safety and optimal dose finding.

Current treatments for Tay-Sachs disease (TSD) are largely supportive, focusing on symptom management rather than modifying the disease course [2]. Investigative therapies aim to slow neurodegeneration, but few have shown meaningful impact on cognitive outcomes or survival. One such therapy, Miglustat, an iminosugar that inhibits glycosphingolipid synthesis, has garnered interest due to its ability to reduce ganglioside accumulation in the brain [3]. Originally developed for Gaucher disease, Miglustat has shown moderate benefit in Niemann-Pick disease type C, another lysosomal storage disorder. In TSD, Miglustat’s effects have been inconsistent across disease types. For infantile-onset, Type 1 TSD, ongoing studies such as the “Effects of Miglustat Therapy on Infantile Type of Sandhoff and Tay-Sachs Diseases (EMTISTD)” trial hopes to recruit 30 participants. While Miglustat does cross the BBB, achieving sufficient CNS concentrations to impact neurodegeneration remains an unanswered question (ClinicalTrials.gov, NCT03822013). Miglustat has shown some benefit in older subjects (Type 2 and Type 3 TSD) by slowing neurological decline, but its impact on disease progression remains modest at best. Based on available data it seems unlikely that Miglustat, as monotherapy, would constitute optimal therapy for young children with Type 1 TSD.

Ambroxol, a pharmacological chaperone known for its mucolytic properties, has shown potential in promoting β-hexosaminidase activity by reducing misfolded protein and enhancing lysosomal function [4,12]. Ambroxol’s action in stabilizing β-hexosaminidase makes it a candidate for treatment of Type 1 TSD, potentially reducing GM2 ganglioside buildup through enhanced lysosomal activity [4,12]. The rationale for combining Miglustat and Ambroxol lies in their complementary mechanisms of action: Miglustat aims to reduce substrate buildup, while Ambroxol could enhance lysosomal activity, collectively supporting neuronal health in TSD [5].

Recent advances in gene therapy for TSD have also shown promise, with early clinical trials indicating potential in increasing enzyme activity and slowing disease progression [9]. However, gene therapy remains experimental, and continued research is needed to establish its safety and long-term efficacy, reinforcing the need for alternative therapeutic options like Miglustat and Ambroxol.

This AI guided virtual study explores the therapeutic potential of combining Miglustat and Ambroxol for treating Type 1 TSD. Given the high lethality of Type 1 TSD before age 10, the study employs a virtual drug trial model focused on young children from birth through age 10. Virtual drug trials offer a promising approach to evaluating treatment efficacy and optimizing dosing in rare diseases like TSD, where patient populations are limited [6,10,11]. Using aiHumanoid simulations, this study aims to assess the impact of low and medium-dose combination therapy on cognitive function and disease progression, providing insights that may guide future clinical strategies for TSD management.

## Methods

### Updating and Validation of the aiHumanoid Simulations (v8.4.4)

The previous version 8.4.3 of the aiHumanoid [13] underwent revisions to v8.4.4. The main differences are that the newest version integrates updated simulations for specific HEXA associated LOF mutations and an updated subsystem for the diagnosis of TSD in young children. As before, the number of integrated organoid simulations remains at 21. To ensure biological fidelity, variability in simulated cohorts was introduced based on population genetic, environmental, and age-specific factors, reflecting real-world diversity in disease progression. The literature validation of the WT and Type 1TSD aiHumanoid simulations employed the same approach used in previous versions to create the updated simulations comprising v8.4.4. [13].

To confirm a diagnosis of Type 1 TSD in the affected aiHumanoid simulated children, a list of 22 genotypic and phenotypic features, was assembled from the peer reviewed literature for evaluation and are presented in Appendix A. All features were statistically significantly different from controls for multiple age matched cohorts and regarding the specific mutation. The present statistical analysis employed a combination of the nonparametric Wilcoxon signed rank test and the Cliff’s delta effect size estimates.

## Study Design

This virtual drug trial leverages aiHumanoid simulations to evaluate the efficacy of Miglustat or Ambroxol, or their combination in treating cognitive decline and other symptoms in children with Type 1 Tay-Sachs Disease (TSD) [2,3]. The trial includes simulated ages from birth to 10 years, with treatment initiation from birth and dose escalation across four treatment levels to explore optimal therapeutic impacts on cognitive function and disease progression. The study includes four arms: Placebo, Miglustat Monotherapy, Ambroxol Monotherapy, and Combination Therapy with Miglustat plus Ambroxol.

## Treatment and Dosage

Each treatment is administered according to an escalating dose schedule to assess tolerability and dose-response effects:

- Initial Dose level: 10% of the Maximum Tolerated Dose (MTD)
- Second Dose level: 20% of MTD
- Third Dose level: 33.3% of MTD
- Fourth Dose level: 50% of MTD

The 50% MTD was selected as the upper dose limit to reduce risks in young populations, balancing efficacy and safety considerations [14]. Dose escalation occurs based on age cohorts to capture the progression of disease impact across early developmental stages in TSD.

Outcomes are evaluated at simulated age intervals of 0-1 years, 2-3 years, 5 years, and 10 years to monitor cumulative treatment effects over time.

### aiHumanoid Simulation Model

The aiHumanoid simulation model replicates a virtual cohort of 25 children in each treatment arm, mimicking biological and physiological characteristics of children with Type 1 TSD. aiHumanoid’s adaptive learning capabilities incorporate real-world TSD progression data and cognitive decline parameters to accurately simulate disease course and intervention effects. The aiHumanoid simulations adjust for variability in subject profiles and drug responses, incorporating stochastic factors relevant to genetic, environmental, and developmental influences on TSD progression [15a,15b].

### Inclusion and Exclusion Criteria

#### Inclusion Criteria

- Simulated patients diagnosed with Type 1 Tay-Sachs Disease (TSD) based on genetic profiles consistent with HEXA gene mutation-induced β-hexosaminidase A deficiency.
- Age at simulation beginning at birth.
- Predicted cognitive and motor decline consistent with Type 1 TSD, with high lethality projected before age 10 years in untreated scenarios.
- Absence of comorbid conditions that could significantly alter the disease trajectory.

#### Exclusion Criteria

- Simulated patients with Type 2 or juvenile forms of TSD, as disease progression and response to treatment may differ.
- Presence of any additional neurodegenerative conditions not directly related to TSD, which could confound treatment outcomes.
- Predicted early mortality within the first 2 years of life based on pre-existing severe neurodegenerative characteristics, to ensure all simulations meet the study’s longitudinal criteria.

### Diagnostic and Treatment Evaluation Features relevant to Type 1 TSD

This AI guided trial evaluates several features, including cognitive and neurodevelopmental metrics, lysosomal and genetic indicators, neurological health, motor and muscle function, quality of life metrics, sensory symptoms, and protein stability markers, such as HEXA levels and lysosomal function [16]. This framework allows the simulation to assess both genotypic markers and phenotypic features, enabling detailed comparisons across placebo, monotherapy, and combination therapy groups (see Appendix A for details).

### Outcome Measures

#### Primary Outcome

- Cognitive Decline Rate: Assessed through cognitive performance scores at each age interval. Decline is measured using a literature derived and standardized cognitive assessment scale embedded in the aiHumanoid simulation, with results compared across treatment arms and dose levels [17].

#### Secondary Outcomes

- Lysosomal Dysfunction
- Motor Function Decline Rate: Assessed via simulated motor skills and task completion rates, compared longitudinally across age cohorts.
- Organ System and General Toxicity Profile: Tracking simulated adverse events across 11 organoid simulations to assess tolerability and safety of combination therapy at maximum dose (50% of MTD).

### Statistical Analysis

The Null hypothesis states that there are no statistically significant differences or at least medium Cliff’s delta effect sizes for the treatment groups compared to the age matched controls treated with placebo or Miglustat alone vs treatment with Miglustat plus Ambroxol.

Outcome measures are analyzed for each treatment arm and dose level, with comparisons made to the placebo group for monotherapies or Miglustat alone for the combination groups using non-parametric methods appropriate for small sample sizes:

- Cognitive Decline and Motor Function: The Wilcoxon Signed-Rank test is used to compare paired differences between treatment arms. Cliff’s delta is calculated to assess effect size, with Hedges corrections for small samples applied to ensure robust analysis [13,18,19].
- Dose-Response Relationship: For each treatment, the relationship between dose levels and cognitive/motor function outcomes is evaluated using the Wilcoxon Signed-Rank test across dose increments, with Cliff’s delta providing effect size [20].
- Healthspan/Longevity: Analyzed using the Wilcoxon Sign Rank test and Cliff’s delta for effectiveness in virtual trials with controlled variability.
- Adverse Events: Risks across dose levels and treatment arms are evaluated with the Wilcoxon Signed-Rank test and Cliff’s delta.

All statistical tests use a significance level of p< 0.05, with Bonferroni corrections applied to account for multiple comparisons (<0.0023 for TSD features and <0.0042 for toxicity estimates) [21,22,23].

## Results

### 1. Miglustat vs Placebo at 10% of MTD

Current therapeutic options Type 1 Tay-Sachs Disease (TSD) are limited, primarily targeting symptom management rather than disease modification. Miglustat, a substrate reduction therapy, has shown potential in preclinical studies for mitigating certain pathological features of TSD. In this virtual clinical trial, we evaluated the impact of Miglustat, administered at 10% of the maximum tolerated dose (MTD) on 22 clinical features associated with TSD across four age groups, focusing on both neuroprotective and disease-modifying effects.

This analysis leverages Cliff’s delta effect sizes to quantify Miglustat’s impact relative to a placebo, with positive values indicating an effect favoring Miglustat and negative values (for features with adverse connotations) also representing a beneficial effect. The results offer insight into the therapeutic potential of Miglustat in altering disease progression and improving quality of life for individuals with Type 1 TSD.

See Appendix B-Table 1: Effect of Miglustat at 10% Maximum Tolerated Dose on Clinical Outcomes in Type 1 Tay-Sachs Disease (TSD)

**Table 1:**
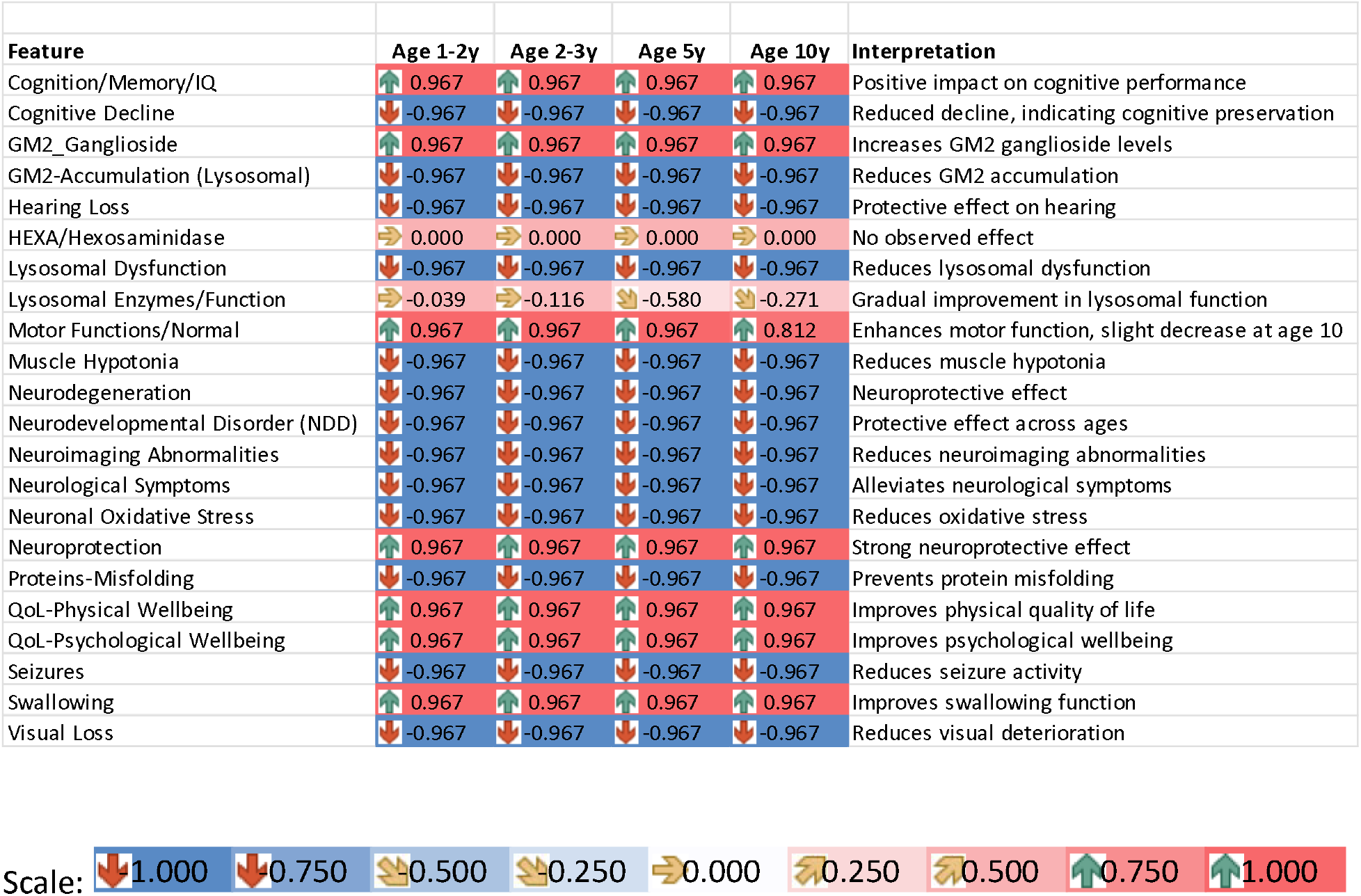
Effect of Miglustat Alone on 22 Disease Features of Type 1 Tay-Sachs Disease at 10% of Maximum Tolerated Dose (MTD)

#### Statistical Test (Wilcoxon Signed-Rank Test)

All corrected p values for comparisons for 21 of the 22 disease features are significant at the p<0.0023 level. The one exception is for HEXA protein levels indicating that the underlying LOF mutation is unaffected by treatment with Miglustat.

#### Summary

In this virtual drug trial, Miglustat at 10% of MTD demonstrated broad beneficial effects across key features, including Cognition/Memory, Motor Function, and Quality of Life (both physical and psychological). Negative Cliff’s delta values on features with adverse implications—such as Cognitive Decline, Neurodegeneration, and Visual Loss—indicate a positive impact for Miglustat, showing potential in reducing the progression of these disease features.

Notably, while Miglustat increased GM2 ganglioside levels, it demonstrated a beneficial reduction in GM2 accumulation within lysosomes. Additionally, a variably negative impact on Lysosomal Enzymes/Function was observed. These findings suggest that Miglustat may offer meaningful symptomatic improvement and neuroprotection in Type 1 TSD, highlighting its potential as a candidate for further investigation at this dose level.

### 2. Ambroxol vs Placebo at 10% of MTD

Ambroxol, is a pharmacological chaperone, which has been studied for its potential to enhance lysosomal function and reduce GM2 ganglioside accumulation in TSD. In this virtual drug trial, we assessed the effects of Ambroxol at 10% of the maximum tolerated dose (MTD) across 22 clinical features and four age cohorts, using Cliff’s delta effect sizes to compare its efficacy to placebo. Positive values indicate a beneficial effect for Ambroxol, while negative values on features with adverse connotations (e.g., Cognitive Decline, Neurodegeneration) also represent positive effects.

See Appendix B-Table 2: Effect of Ambroxol at 10% Maximum Tolerated Dose on Clinical Outcomes in Type 1 Tay-Sachs Disease (TSD)

**Table 2:** Effect of Ambroxol at 10% Maximum Tolerated Dose on Clinical Outcomes in Type 1 Tay-Sachs Disease (TSD)

| Feature | Age 1-2y | Age 2-3y | Age 5y | Age 10y | Interpretation |
| --- | --- | --- | --- | --- | --- |
| Cognition/Memory/IQ | ↑ 0.967 | ↑ 0.967 | ↑ 0.967 | ↑ 0.967 | Positive impact on cognitive performance |
| Cognitive Decline | ↓ -0.967 | ↓ -0.967 | ↓ -0.967 | ↓ -0.967 | Reduced decline, indicating cognitive preservation |
| GM2_Ganglioside | ↑ 0.967 | ↑ 0.967 | ↑ 0.890 | ↑ 0.890 | Increases GM2 ganglioside levels |
| GM2-Accumulation (Lysosomal) | ↓ -0.967 | ↓ -0.967 | ↓ -0.967 | ↓ -0.967 | Reduces GM2 accumulation |
| Hearing Loss | ↓ -0.967 | ↓ -0.967 | ↓ -0.967 | ↓ -0.967 | Protective effect on hearing |
| HEXA/Hexosaminidase | → 0.000 | → 0.000 | → 0.000 | → 0.000 | No observed effect |
| Lysosomal Dysfunction | ↓ -0.967 | ↓ -0.967 | ↓ -0.967 | ↓ -0.967 | Reduces lysosomal dysfunction |
| Lysosomal Enzymes/Function | → 0.039 | → -0.039 | → 0.039 | ↑ 0.735 | Increased lysosomal enzyme activity seen at age 10 years |
| Motor Functions/Normal | ↑ 0.967 | ↑ 0.967 | ↑ 0.967 | ↑ 0.812 | Enhanced motor function |
| Muscle Hypotonia | ↓ -0.967 | ↓ -0.967 | ↓ -0.967 | ↓ -0.967 | Reduces muscle hypotonia |
| Neurodegeneration | ↓ -0.967 | ↓ -0.967 | ↓ -0.967 | ↓ -0.967 | Neuroprotective effect |
| Neurodevelopmental Disorder (NDD) | ↓ -0.967 | ↓ -0.967 | ↓ -0.967 | ↓ -0.967 | Protective effect across ages |
| Neuroimaging Abnormalities | ↓ -0.967 | ↓ -0.967 | ↓ -0.967 | ↓ -0.967 | Reduces neuroimaging abnormalities |
| Neurological Symptoms | ↓ -0.967 | ↓ -0.967 | ↓ -0.967 | ↓ -0.967 | Alleviates neurological symptoms |
| Neuronal Oxidative Stress | ↓ -0.967 | ↓ -0.967 | ↓ -0.967 | ↓ -0.967 | Reduces oxidative stress |
| Neuroprotection | ↑ 0.967 | ↑ 0.967 | ↑ 0.967 | ↑ 0.967 | Strong neuroprotective effect |
| Proteins-Misfolding | ↓ -0.967 | ↓ -0.967 | ↓ -0.967 | ↓ -0.967 | Prevents protein misfolding |
| QoL-Physical Wellbeing | ↑ 0.967 | ↑ 0.967 | ↑ 0.967 | ↑ 0.967 | Improved physical quality of life |
| QoL-Psychological Wellbeing | ↑ 0.967 | ↑ 0.967 | ↑ 0.967 | ↑ 0.967 | Improved psychological wellbeing |
| Seizures | ↓ -0.967 | ↓ -0.967 | ↓ -0.967 | ↓ -0.967 | Reduces seizure activity |
| Swallowing | ↑ 0.967 | ↑ 0.967 | ↑ 0.967 | ↑ 0.967 | Improves swallowing function |
| Visual Loss | ↓ -0.967 | ↓ -0.967 | ↓ -0.967 | ↓ -0.967 | Reduces visual deterioration |

#### Statistical Test (Wilcoxon Signed-Rank Test)

All corrected p values for comparisons for 21 of the 22 disease features are significant at the p<0.0023 level. The one exception is for HEXA protein levels indicating that the underlying LOF mutation in unaffected by treatment with Ambroxol.

#### Summary

Ambroxol at 10% MTD displayed beneficial effects across a wide range of clinical features in Type 1 TSD. Positive Cliff’s delta values were observed for Cognition/Memory, Motor Function, and Quality of Life measures, suggesting that Ambroxol could enhance these outcomes. Negative effects on adverse features—such as Cognitive Decline, Neurodegeneration, and Visual Loss—indicate a protective effect, consistent across all ages studied.

Ambroxol increased GM2 ganglioside levels but showed a favorable impact on reducing GM2 accumulation within lysosomes. Additionally, there was a significant improvement in Lysosomal Enzymes/Function, particularly at age 10 years, suggesting a cumulative effect on lysosomal activity. Overall, these findings underscore Ambroxol’s potential to mitigate neurodegeneration and improve quality-of-life metrics in Type 1 TSD, warranting further exploration at this dosage level.

### 3. Miglustat Plus Ambroxol vs Miglustat alone at 10% of MTD

Current therapeutic interventions are limited, prompting investigation into combination therapies to enhance treatment efficacy. In this virtual trial, we evaluated the combination of Miglustat and Ambroxol, both administered at 10% of the maximum tolerated dose (MTD), compared to Miglustat alone. This analysis uses Cliff’s delta effect sizes to quantify the effect of the combination therapy relative to Miglustat monotherapy across 22 clinical features. Positive values favor the combination, while negative values (for adverse features) indicate a positive effect for the combination as well.

See Appendix B-Table 3: Effect of Miglustat plus Ambroxol at 10% MTD Compared to Miglustat Alone on Clinical Outcomes in Type 1 Tay-Sachs Disease (TSD)

**Table 3:** Effect of Miglustat+Abroxol vs Miglutat alone at 10% of Maximum Tolerated Dose (MTD)

| Feature | Age 1-2y | Age 2-3y | Age 5y | Age 10y | Interpretation: Combination vs Miglustat alone |
| --- | --- | --- | --- | --- | --- |
| Cognition/Memory/IQ | ➡ 0.000 | ➡ 0.000 | ➡ 0.000 | ➡ 0.000 | No additional impact on cognition |
| Cognitive Decline | ➡ -0.039 | ➡ -0.425 | ➡ -0.503 | ➡ -0.619 | Progressive cognitive preservation with age |
| GM2_Ganglioside | ➡ -0.967 | ➡ -0.967 | ➡ -0.967 | ➡ -0.271 | Reduced GM2 levels, notably at younger ages |
| GM2-Accumulation (Lysosomal) | ➡ -0.812 | ➡ -0.735 | ➡ 0.658 | ➡ 0.967 | Improvement at younger ages, increased at older ages |
| Hearing Loss | ➡ 0.890 | ➡ 0.890 | ➡ 0.967 | ➡ 0.928 | Hearing preservation or improvement |
| HEXA/Hexosaminidase | ➡ 0.000 | ➡ 0.000 | ➡ 0.000 | ➡ 0.000 | No observed effect |
| Lysosomal Dysfunction | ➡ 0.967 | ➡ 0.967 | ➡ 0.967 | ➡ 0.967 | Consistent reduction in lysosomal function |
| Lysosomal Enzymes/Function | ➡ 0.812 | ➡ 0.812 | ➡ 0.812 | ➡ 0.735 | Enhanced enzyme activity with slight decrease at age 10y |
| Motor Functions/Normal | ➡ 0.967 | ➡ 0.967 | ➡ 0.967 | ➡ 0.967 | Strong positive impact on motor function |
| Muscle Hypotonia | ➡ -0.967 | ➡ -0.967 | ➡ -0.967 | ➡ -0.967 | Reduction in muscle hypotonia |
| Neurodegeneration | ➡ -0.967 | ➡ -0.967 | ➡ -0.967 | ➡ -0.967 | Neuroprotective effect |
| Neurodevelopmental Disorder (NDD) | ➡ -0.967 | ➡ -0.967 | ➡ -0.967 | ➡ -0.967 | Protective effect across ages |
| Neuroimaging Abnormalities | ➡ -0.967 | ➡ -0.967 | ➡ -0.967 | ➡ -0.967 | Reduction in neuroimaging abnormalities |
| Neurological Symptoms | ➡ -0.967 | ➡ -0.967 | ➡ -0.967 | ➡ -0.967 | Alleviation of neurological symptoms |
| Neuronal Oxidative Stress | ➡ -0.967 | ➡ -0.967 | ➡ -0.967 | ➡ -0.967 | Reduction in oxidative stress |
| Neuroprotection | ➡ -0.967 | ➡ -0.967 | ➡ -0.967 | ➡ -0.967 | Consistent neuroprotection |
| Proteins-Misfolding | ➡ -0.425 | ➡ -0.967 | ➡ -0.890 | ➡ -0.890 | Reduced protein misfolding |
| QoL-Physical Wellbeing | ➡ 0.851 | ➡ 0.542 | ➡ 0.619 | ➡ 0.890 | Improvement in physical quality of life |
| QoL-Psychological Wellbeing | ➡ 0.967 | ➡ 0.967 | ➡ 0.967 | ➡ 0.928 | Improvement in psychological wellbeing |
| Seizures | ➡ -0.967 | ➡ -0.967 | ➡ -0.928 | ➡ -0.890 | Reduction in seizure frequency |
| Swallowing | ➡ 0.967 | ➡ 0.967 | ➡ 0.967 | ➡ 0.967 | Improved swallowing function |
| Visual Loss | ➡ -0.967 | ➡ -0.967 | ➡ -0.967 | ➡ -0.967 | Reduction in visual deterioration |

Scale: ➡ 1.000 ➡ 0.750 ➡ 0.500 ➡ 0.250 ➡ 0.000 ➡ 0.250 ➡ 0.500 ➡ 0.750 ➡ 1.000

#### Statistical Test (Wilcoxon Signed-Rank Test)

The corrected p values for comparisons for 19 of the 22 disease features are significant at the p<0.0023 level. Cognitive function and cognitive decline are not significantly different between Miglustat plus Ambroxol vs Miglustat alone (p>0.0023). The other exception is once again for HEXA protein levels indicating that the underlying LOF mutation is unaffected by treatment with Miglustat plus Ambroxol.

#### Summary

This evaluation of Miglustat combined with Ambroxol at 10% MTD demonstrated additional therapeutic benefits over Miglustat alone in several key clinical areas. The combination therapy produced consistent improvement in Cognitive Decline, Neurodegeneration, and Visual Loss, with a progressively stronger effect on cognitive preservation as the cohorts age. Additionally, the combination therapy significantly reduced GM2 ganglioside levels across all ages, indicating effective substrate reduction, particularly at younger ages. However, there was an increase in GM2 accumulation in lysosomes at older ages, suggesting a potential age-related limitation in efficacy.

Ambroxol contributed significantly to improved Lysosomal Dysfunction and Lysosomal Enzymes/Function, with a consistent effect across age groups, although enzyme function showed a slight reduction by age 10. Motor Functions and Muscle Hypotonia also benefited from the combination therapy, with strong positive impacts across all ages. Quality of Life (QoL) measures, both physical and psychological, were notably improved, especially in older cohorts.

Overall, these data indicate that the combination of Miglustat plus Ambroxol offers a promising therapeutic benefit, particularly for younger patients, with enhancements in both neuroprotection and quality of life compared to Miglustat alone. Further studies are warranted to assess the age-related efficacy trends and potential optimizations for older children with Type 1 TSD.

### 4. Miglustat vs Placebo at 20% of MTD

With this analysis we evaluate the effect of Miglustat at 20% of the maximum tolerated dose (MTD) compared to Placebo across 22 clinical features, using Cliff’s delta effect sizes. Positive values favor Miglustat, while negative values (for negatively connoted features) indicate a positive effect for Miglustat by reducing the severity of those adverse features.

See Appendix B-Table 4: Effect of Miglustat at 20% MTD Compared to Placebo on Clinical Outcomes in Type 1 Tay-Sachs Disease (TSD)

**Table 4:** Effect of Miglustat at 20% MTD Compared to Placebo on Clinical Outcomes in Type 1 Tay-Sachs Disease (TSD)

| Feature | Age 1-2y | Age 2-3y | Age 5y | Age 10y | Interpretation |
| --- | --- | --- | --- | --- | --- |
| Cognition/Memory/IQ | ➡ 0.967 | ➡ 0.967 | ➡ 0.967 | ➡ 0.967 | Positive impact on cognitive function |
| Cognitive Decline | ➡ -0.967 | ➡ -0.967 | ➡ -0.967 | ➡ -0.967 | Cognitive preservation |
| GM2_Ganglioside | ➡ 0.967 | ➡ 0.967 | ➡ 0.967 | ➡ 0.967 | Increased GM2 levels |
| GM2-Accumulation (Lysosomal) | ➡ -0.967 | ➡ -0.967 | ➡ -0.967 | ➡ -0.967 | Reduced lysosomal GM2 accumulation |
| Hearing Loss | ➡ -0.967 | ➡ -0.967 | ➡ -0.967 | ➡ -0.967 | Protective effect on hearing |
| HEXA/Hexosaminidase | ➡ 0.000 | ➡ 0.000 | ➡ 0.000 | ➡ 0.000 | No observed effect |
| Lysosomal Dysfunction | ➡ -0.967 | ➡ -0.967 | ➡ -0.967 | ➡ -0.967 | Reduction in lysosomal dysfunction |
| Lysosomal Enzymes/Function | ➡ -0.735 | ➡ -0.967 | ➡ -0.890 | ➡ -0.193 | Decline in enzyme function, some improvement with age |
| Motor Functions/Normal | ➡ 0.967 | ➡ 0.967 | ➡ 0.967 | ➡ 0.812 | Positive impact on motor function |
| Muscle Hypotonia | ➡ -0.967 | ➡ -0.967 | ➡ -0.967 | ➡ -0.967 | Reduced muscle hypotonia |
| Neurodegeneration | ➡ -0.967 | ➡ -0.967 | ➡ -0.967 | ➡ -0.967 | Neuroprotective effect |
| Neurodevelopmental Disorder (NDD) | ➡ -0.967 | ➡ -0.967 | ➡ -0.967 | ➡ -0.967 | Consistent reduction in neurodevelopmental symptoms |
| Neuroimaging Abnormalities | ➡ -0.967 | ➡ -0.967 | ➡ -0.967 | ➡ -0.967 | Reduced abnormalities in neuroimaging |
| Neurological Symptoms | ➡ -0.967 | ➡ -0.967 | ➡ -0.967 | ➡ -0.967 | Alleviation of neurological symptoms |
| Neuronal Oxidative Stress | ➡ -0.967 | ➡ -0.967 | ➡ -0.967 | ➡ -0.967 | Reduction in oxidative stress |
| Neuroprotection | ➡ 0.967 | ➡ 0.967 | ➡ 0.967 | ➡ 0.967 | Large neuroprotective effect |
| Proteins-Misfolding | ➡ -0.967 | ➡ -0.967 | ➡ -0.967 | ➡ -0.967 | Reduced protein misfolding |
| QoL-Physical Wellbeing | ➡ 0.967 | ➡ 0.967 | ➡ 0.967 | ➡ 0.967 | Improvement in physical quality of life |
| QoL-Psychological Wellbeing | ➡ 0.967 | ➡ 0.967 | ➡ 0.967 | ➡ 0.967 | Improved psychological wellbeing |
| Seizures | ➡ -0.967 | ➡ -0.967 | ➡ -0.967 | ➡ -0.967 | Reduction in seizure activity |
| Swallowing | ➡ 0.967 | ➡ 0.967 | ➡ 0.967 | ➡ 0.967 | Improved swallowing function |
| Visual Loss | ➡ -0.967 | ➡ -0.967 | ➡ -0.967 | ➡ -0.967 | Reduced visual deterioration |

#### Statistical Test (Wilcoxon Signed-Rank Test)

All corrected p values for comparisons for 20 of the 22 disease features are significant at the p<0.0023 level. At age 10 years the difference between Lysosomal enzyme function is not statistically significant (p>0.0023). The only other exception is for HEXA protein levels indicating once again that the underlying LOF mutation is unaffected by treatment with Miglustat.

## Summary

In this part of the virtual drug trial, Miglustat at 20% MTD demonstrated significant positive effects on multiple clinical features in Type 1 Tay-Sachs Disease (TSD) compared to Placebo. Miglustat showed a beneficial impact on Cognition/Memory, Motor Function, and Quality of Life (both physical and psychological), maintaining high positive effect sizes across all ages. Additionally, negative Cliff’s delta values for features with negative connotations—such as Cognitive Decline, Neurodegeneration, and Visual Loss—highlight the neuroprotective potential of Miglustat in slowing disease progression.

Miglustat also reduced GM2 accumulation in lysosomes across all age groups, although it was associated with increased GM2 ganglioside levels, which may indicate substrate load effects. A consistent improvement was noted in Lysosomal Enzymes/Function at younger ages, although this effect was less pronounced at age 10, suggesting a potential decrease in efficacy with age.

Overall, the data indicate that Miglustat at 20% MTD could offer substantial benefits in managing Type 1 TSD symptoms, especially for younger patients, providing both symptomatic relief and neuroprotective effects. Further investigation into age-specific dosing and long-term outcomes could help optimize its therapeutic application.

### 5. Ambroxol vs Placebo at 20% of MTD

Here we evaluate the effects of Ambroxol at 20% of (MTD) compared to Placebo across 22 clinical features, using Cliff’s delta effect sizes. Positive values favor Ambroxol, while negative values (for negatively connoted features) also indicate a positive effect by reducing the severity of those adverse features.

See Appendix B-Table 5: Effect of Ambroxol at 20% MTD Compared to Placebo on Clinical Outcomes in Type 1 Tay-Sachs Disease (TSD)

**Table 5:**
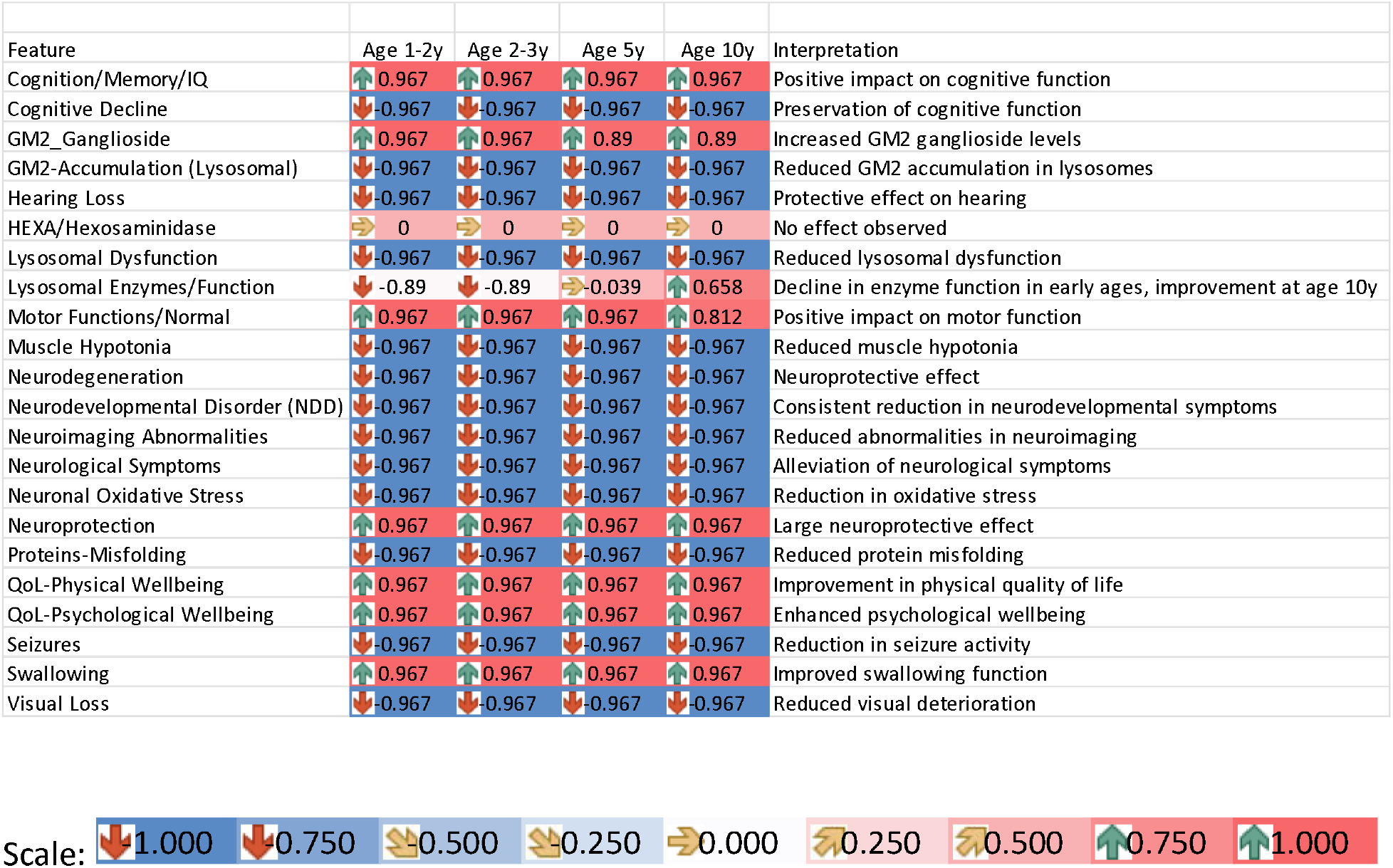
Effect of Ambroxol at 20% MTD Compared to Placebo on Clinical Outcomes in Type 1 Tay-Sachs Disease (TSD)

#### Statistical Test (Wilcoxon Signed-Rank Test)

All corrected p values for comparisons for 20 of the 22 disease features are significant at the p<0.0023 level. At ages 5 and 10 years the difference between Lysosomal enzyme function is not statistically significant (p>0.0023). The only other exception is for HEXA protein levels indicating that the underlying LOF mutation is unaffected by treatment with Ambroxol.

#### Summary

In this virtual drug trial, Ambroxol at 20% MTD demonstrated a range of beneficial effects on multiple clinical features in Type 1 Tay-Sachs Disease (TSD) compared to Placebo. Ambroxol showed a positive impact on Cognition/Memory, Motor Function, and Quality of Life measures (both physical and psychological) across all ages. Negative Cliff’s delta values for adverse features—such as Cognitive Decline, Neurodegeneration, and Visual Loss—highlight the neuroprotective potential of Ambroxol in slowing disease progression.

For Lysosomal Enzymes/Function, the data indicate a decline in enzyme function at younger ages (1-5 years), as represented by negative Cliff’s delta values. However, there is an observed improvement by age 10, suggesting a delayed positive effect of Ambroxol on lysosomal enzyme function over time. This delayed response could imply an adaptive or cumulative benefit that becomes more apparent in older children.

The findings suggest that Ambroxol at 20% MTD has a favorable impact on several clinical aspects of Type 1 TSD, including cognitive and motor functions and neuroprotection. However, the initial decline in Lysosomal Enzymes/Function at younger ages requires clinical validation, as it may indicate a transient adverse effect that improves with continued treatment. Further studies are recommended to explore the age-dependent dynamics of Ambroxol’s impact on lysosomal function, to optimize dosing strategies that maximize therapeutic benefits while minimizing early childhood adverse effects.

### 6. Miglustat Plus Ambroxol vs Miglustat alone at 20% of MTD

This analysis examines the combined effects of Miglustat and Ambroxol, each at 20% of their maximum tolerated doses (MTD), compared to Miglustat monotherapy also at 29% MTD. Cliff’s delta effect sizes are used to measure the efficacy of the combination therapy across 22 clinical features. Positive values favor the combination therapy, while negative values (for negatively connoted features) indicate a beneficial effect from the combination therapy.

See Appendix B-Table 6: Effect of Miglustat plus Ambroxol at 20% MTD Compared to Miglustat Alone on Clinical Outcomes in Type 1 Tay-Sachs Disease (TSD)

**Table 6:**
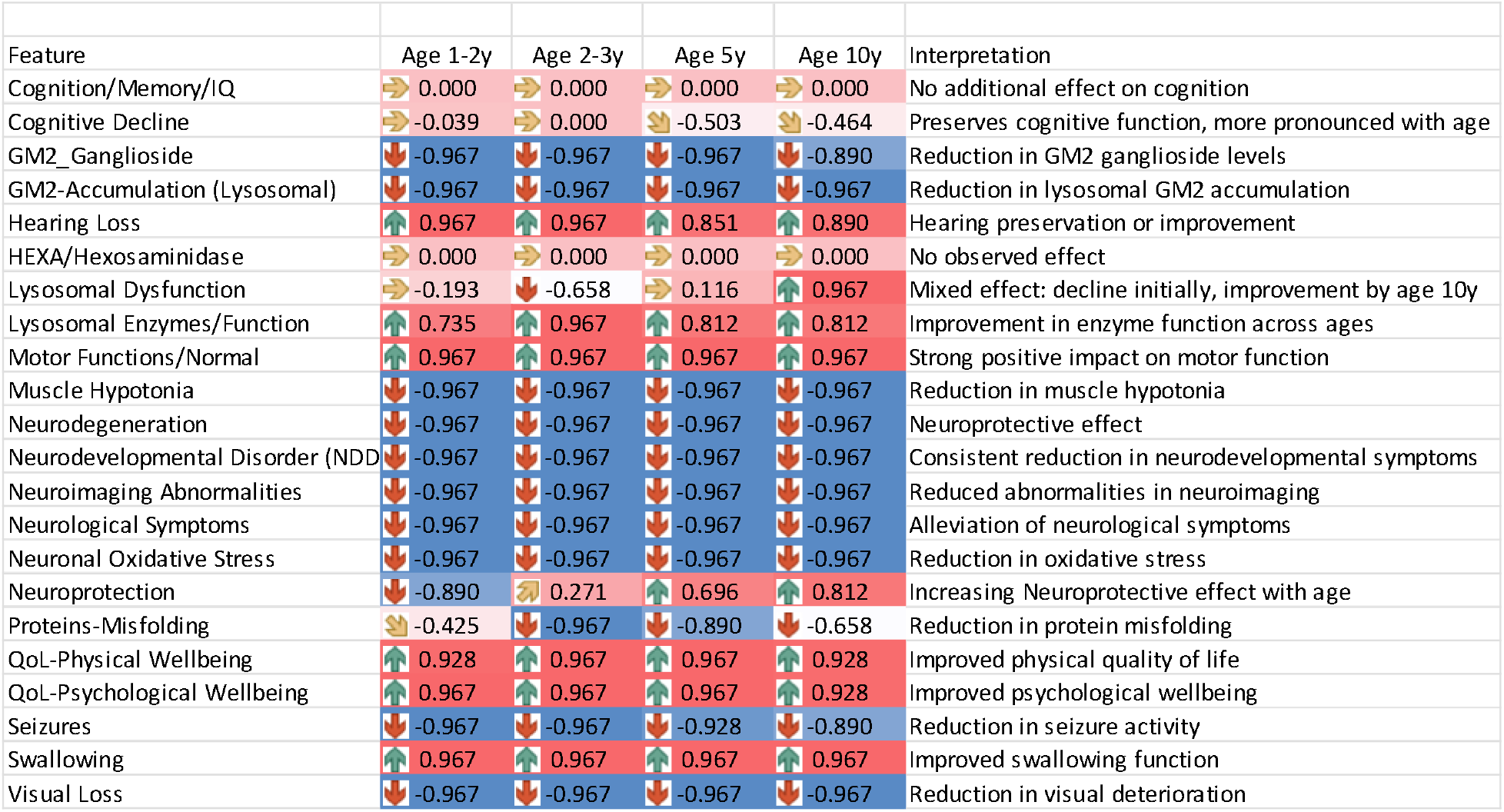
Effect of Miglustat + Ambroxol at 20% MTD Compared to Miglustat Alone on Clinical Outcomes in Type 1 Tay-Sachs Disease (TSD)

#### Statistical Test (Wilcoxon Signed-Rank Test)

At birth to 1 years the differences between Cognition/Memory/IQ, Cognitive Decline, HEXA, and Lysosomal dysfunction are not statistically significant. By age 2-3years Lysosomal dysfunction improves but Cognition/Memory/IQ, Cognitive Decline, and HEXA differences fail to achieve statistical significance. At age 5 years differences in Cognition/Memory/IQ, Lysosomal Dysfunction and HEXA differences fail to achieve statistical significance. By age 10 years all features except Cognitive Function and HEXA achieved statistical significance.

#### Summary

In this virtual clinical trial, the combination of Miglustat and Ambroxol at 20% MTD demonstrated several beneficial effects across key clinical features in Type 1 Tay-Sachs Disease (TSD) compared to Miglustat alone. Notable benefits of the combination therapy were observed in Cognitive Decline and Neurodegeneration, with negative values suggesting preservation of cognitive and neurological function, especially in older children.

GM2 Ganglioside and GM2-Accumulation (Lysosomal) levels show consistent reductions across all age groups, indicating effective substrate reduction with combination therapy. Lysosomal Dysfunction results are mixed, with initial declines in function (1-2 and 2-3 years), followed by improvement by age 10, suggesting an adaptive response with extended therapy.

Lysosomal Enzymes/Function showed consistent improvement across all age groups with positive values, indicating enhanced enzymatic function relative to Miglustat alone. This effect may reflect an additive benefit from Ambroxol on lysosomal activity.

Improvements were also noted in Quality of Life measures (both physical and psychological) and Motor Functions, with the combination therapy providing consistent positive effects across all ages. Neuroprotection was seen to increase progressively with age, suggesting a cumulative benefit over time.

The combination of Miglustat and Ambroxol at 20% MTD offers several potential advantages over Miglustat alone in managing Type 1 TSD. Benefits include enhanced neuroprotection, reduced GM2 ganglioside levels, and improved quality of life, with mixed but overall positive effects on lysosomal function. Further research into the age-related dynamics of lysosomal enzyme function and the potential cumulative effects of combination therapy is recommended to optimize therapeutic strategies for TSD.

### 7. Miglustat vs Placebo at 33.3% of MTD

This analysis evaluates the effects of Miglustat at 33.3% MTD compared to Placebo across 22 clinical features in four age groups (1-2 years, 2-3 years, 5 years, and 10 years) in Type 1 Tay-Sachs Disease (TSD). Each feature’s Cliff’s delta effect size is examined, where positive values indicate a favorable outcome for Miglustat over Placebo. Additionally, we will use the Wilcoxon Signed-Rank Test across features to determine if there are a statistically significant difference (p<0.0023).

See Appendix B-Table 7: Effect of Miglustat at 33.3% MTD Compared to Placebo on Clinical Outcomes in Type 1 Tay-Sachs Disease (TSD)

**Table 7:** Effect of Miglustat at 33.3% MTD Compared to Placebo on Clinical Outcomes in Type 1 Tay-Sachs Disease (TSD)

| Feature | Age 1-2y | Age 2-3y | Age 5y | Age 10y | Interpretation |
| --- | --- | --- | --- | --- | --- |
| Cognition/Memory/IQ | ↑0.967 | ↑0.967 | ↑0.967 | ↑0.967 | Positive impact on cognitive function |
| Cognitive Decline | ↓-0.967 | ↓-0.967 | ↓-0.967 | ↓-0.967 | Preservation of cognitive function |
| GM2_Ganglioside | ⇒-0.039 | ⇒-0.039 | ⇒-0.039 | ⇒0.039 | No apparent difference |
| GM2-Accumulation (Lysosomal) | ↓-0.967 | ↓-0.967 | ↓-0.967 | ↓-0.967 | Reduction in lysosomal GM2 accumulation |
| Hearing Loss | ↓-0.967 | ↓-0.967 | ↓-0.967 | ↓-0.967 | Protective effect on hearing |
| HEXA/Hexosaminidase | ⇒0.000 | ⇒0.000 | ⇒0.000 | ⇒0.000 | No observed effect |
| Lysosomal Dysfunction | ↓-0.967 | ↓-0.967 | ↓-0.967 | ↓-0.967 | Reduction in lysosomal dysfunction |
| Lysosomal Enzymes/Function | ↓-0.967 | ↓-0.967 | ↓-0.967 | ↓-0.967 | Decline in enzyme function |
| Motor Functions/Normal | ↑0.967 | ↑0.967 | ↑0.967 | ↑0.967 | Large positive impact on motor function |
| Muscle Hypotonia | ↓-0.967 | ↓-0.967 | ↓-0.967 | ↓-0.967 | Reduction in muscle hypotonia |
| Neurodegeneration | ↓-0.967 | ↓-0.967 | ↓-0.967 | ↓-0.967 | Neuroprotective effect |
| Neurodevelopmental Disorder (NDD) | ↓-0.967 | ↓-0.967 | ↓-0.967 | ↓-0.967 | Consistent reduction in neurodevelopmental symptoms |
| Neuroimaging Abnormalities | ↓-0.967 | ↓-0.967 | ↓-0.967 | ↓-0.967 | Reduced abnormalities in neuroimaging |
| Neurological Symptoms | ↓-0.967 | ↓-0.967 | ↓-0.967 | ↓-0.967 | Alleviation of neurological symptoms |
| Neuronal Oxidative Stress | ↓-0.967 | ↓-0.967 | ↓-0.967 | ↓-0.967 | Reduction in oxidative stress |
| Neuroprotection | ↑0.967 | ↑0.967 | ↑0.967 | ↑0.967 | Large neuroprotective effect |
| Proteins-Misfolding | ↓-0.967 | ↓-0.967 | ↓-0.967 | ↓-0.967 | Reduction in protein misfolding |
| QoL-Physical Wellbeing | ↑0.967 | ↑0.967 | ↑0.967 | ↑0.967 | Improved physical quality of life |
| QoL-Psychological Wellbeing | ↑0.967 | ↑0.967 | ↑0.967 | ↑0.967 | Improved psychological wellbeing |
| Seizures | ↓-0.967 | ↓-0.967 | ↓-0.967 | ↓-0.967 | Reduction in seizure activity |
| Swallowing | ↑0.967 | ↑0.967 | ↑0.967 | ↑0.967 | Improved swallowing function |
| Visual Loss | ↓-0.967 | ↓-0.967 | ↓-0.967 | ↓-0.967 | Reduction in visual deterioration |

#### Statistical Test (Wilcoxon Signed-Rank Test)

All corrected p values for comparisons for 20 of the 22 disease features are significant at the p<0.0023 level. At all ages there are no statistically significant differences between GM2 Ganglioside levels between Miglustat and Placebo effects. The only other exception is for HEXA protein levels indicating that the underlying LOF mutation is unaffected by treatment with Miglustat.

#### Summary

This analysis of Miglustat at 33.3% MTD demonstrated beneficial effects across several clinical features in Type 1 TSD compared to Placebo:

Consistent Positive Effects: Large positive effect sizes were observed on Cognition/Memory, Motor Function, and Quality of Life (both physical and psychological), with Cliff’s delta values consistently at 0.967 across all age groups.

Reduction in Negative Outcomes: Negative effect sizes for features such as Cognitive Decline, Neurodegeneration, Visual Loss, and Neurodevelopmental Disorders indicate reductions in these adverse outcomes, highlighting a neuroprotective benefit from Miglustat at this dose level.

GM2 Ganglioside: The effect sizes for GM2 Ganglioside are small (−0.039 to +0.039 across ages), indicating trivial differences between Miglustat and Placebo in terms of ganglioside levels. However, a consistent reduction is noted in GM2-Accumulation (Lysosomal), suggesting that the dose helps limit GM2 build-up in lysosomes specifically.

Lysosomal Enzymes/Function: A negative effect size (−0.967 across ages) suggests a decline in enzyme function relative to Placebo, which may warrant further investigation to determine if this dose of Miglustat has unintended effects on lysosomal enzyme activity.

### 8. Ambroxol vs Placebo at 33.3% MTD

This analysis evaluates the effects of Ambroxol at 33.3% MTD compared to Placebo across 22 clinical features in four age groups (1-2 years, 2-3 years, 5 years, and 10 years) in Type 1 Tay-Sachs Disease (TSD). Each feature’s Cliff’s delta effect size is examined, where positive values indicate a favorable outcome for Ambroxol over Placebo. Wilcoxon Signed-Rank Tests with Bonferroni correction were conducted to assess the statistical significance of the differences across features.

See Appendix B-Table 8: Effect of Ambroxol at 33.3% MTD Compared to Placebo on Clinical Outcomes in Type 1 Tay-Sachs Disease (TSD)

**Table 8:**
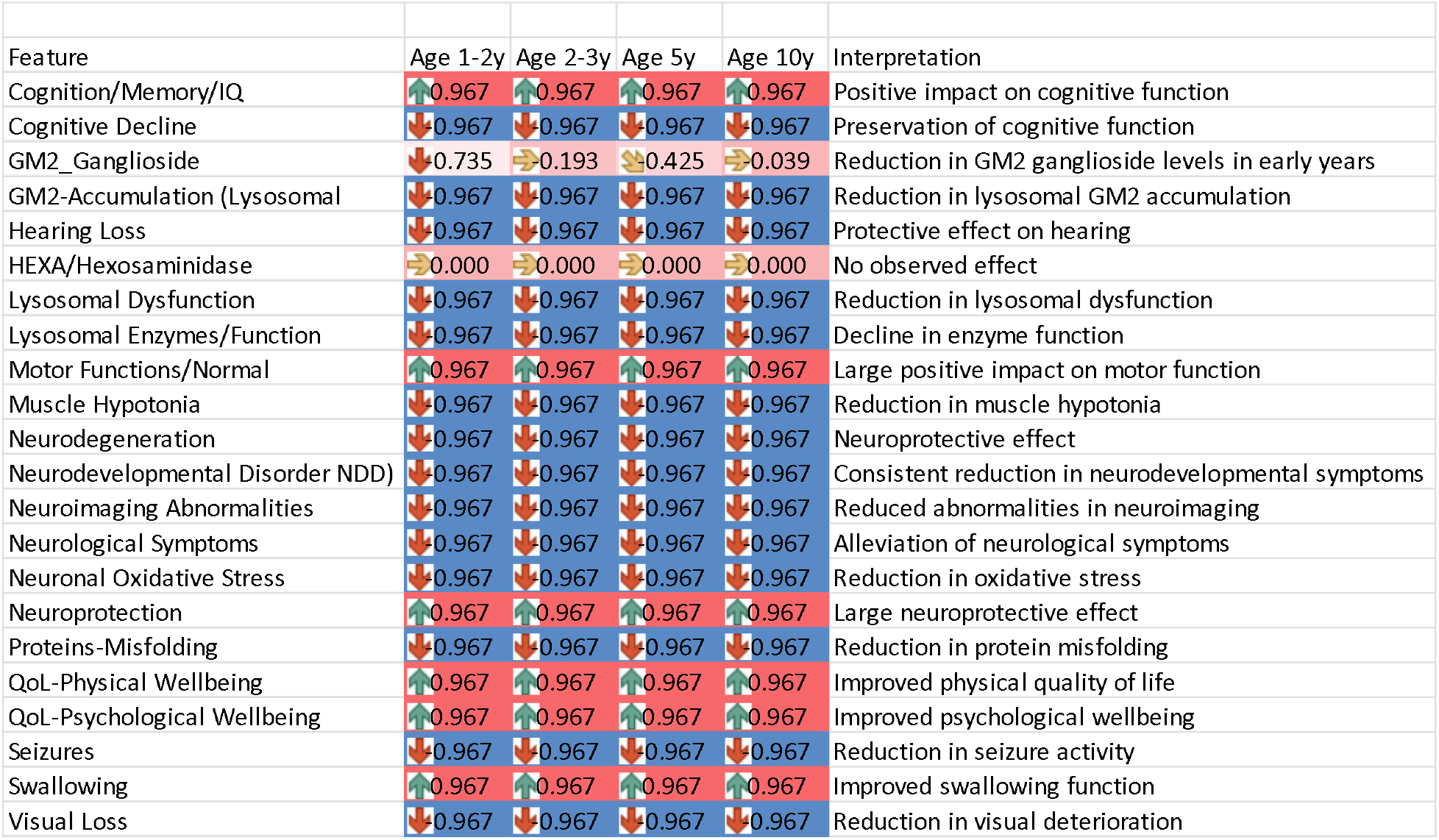
Effect of Ambroxol at 33.3% MTD Compared to Placebo in Type 1 Tay-Sachs Disease (TSD)

| Feature | Age 1-2y | Age 2-3y | Age 5y | Age 10y | Interpretation |
| --- | --- | --- | --- | --- | --- |
| Cognition/Memory/IQ | ↑0.967 | ↑0.967 | ↑0.967 | ↑0.967 | Positive impact on cognitive function |
| Cognitive Decline | ↓0.967 | ↓0.967 | ↓0.967 | ↓0.967 | Preservation of cognitive function |
| GM2_Ganglioside | ↓0.735 | ⇒0.193 | ⇒0.425 | ⇒0.039 | Reduction in GM2 ganglioside levels in early years |
| GM2-Accumulation (Lysosomal) | ↓0.967 | ↓0.967 | ↓0.967 | ↓0.967 | Reduction in lysosomal GM2 accumulation |
| Hearing Loss | ↓0.967 | ↓0.967 | ↓0.967 | ↓0.967 | Protective effect on hearing |
| HEXA/Hexosaminidase | ⇒0.000 | ⇒0.000 | ⇒0.000 | ⇒0.000 | No observed effect |
| Lysosomal Dysfunction | ↓0.967 | ↓0.967 | ↓0.967 | ↓0.967 | Reduction in lysosomal dysfunction |
| Lysosomal Enzymes/Function | ↓0.967 | ↓0.967 | ↓0.967 | ↓0.967 | Decline in enzyme function |
| Motor Functions/Normal | ↑0.967 | ↑0.967 | ↑0.967 | ↑0.967 | Large positive impact on motor function |
| Muscle Hypotonia | ↓0.967 | ↓0.967 | ↓0.967 | ↓0.967 | Reduction in muscle hypotonia |
| Neurodegeneration | ↓0.967 | ↓0.967 | ↓0.967 | ↓0.967 | Neuroprotective effect |
| Neurodevelopmental Disorder (NDD) | ↓0.967 | ↓0.967 | ↓0.967 | ↓0.967 | Consistent reduction in neurodevelopmental symptoms |
| Neuroimaging Abnormalities | ↓0.967 | ↓0.967 | ↓0.967 | ↓0.967 | Reduced abnormalities in neuroimaging |
| Neurological Symptoms | ↓0.967 | ↓0.967 | ↓0.967 | ↓0.967 | Alleviation of neurological symptoms |
| Neuronal Oxidative Stress | ↓0.967 | ↓0.967 | ↓0.967 | ↓0.967 | Reduction in oxidative stress |
| Neuroprotection | ↑0.967 | ↑0.967 | ↑0.967 | ↑0.967 | Large neuroprotective effect |
| Proteins-Misfolding | ↓0.967 | ↓0.967 | ↓0.967 | ↓0.967 | Reduction in protein misfolding |
| QoL-Physical Wellbeing | ↑0.967 | ↑0.967 | ↑0.967 | ↑0.967 | Improved physical quality of life |
| QoL-Psychological Wellbeing | ↑0.967 | ↑0.967 | ↑0.967 | ↑0.967 | Improved psychological wellbeing |
| Seizures | ↓0.967 | ↓0.967 | ↓0.967 | ↓0.967 | Reduction in seizure activity |
| Swallowing | ↑0.967 | ↑0.967 | ↑0.967 | ↑0.967 | Improved swallowing function |
| Visual Loss | ↓0.967 | ↓0.967 | ↓0.967 | ↓0.967 | Reduction in visual deterioration |

By age 1 year there is a decline in GM2 gangliosides (p<0.0023). From age 2-3 years to age 10 years inclusive differences in GM2 Ganglioside levels are no longer different (p>0.0023). Consistent with previous results there were no differences between HEXA levels. All other differences were statistically significant at the p<0.0023 level.

#### Summary

Positive Effects: Significant positive impacts are observed on Cognition/Memory, Motor Functions, and Quality of Life (both physical and psychological), with a consistently large Cliff’s delta of 0.967 across ages.

Reduction in Negative Outcomes: Features such as GM2 Ganglioside, GM2-Accumulation, Neurodegeneration, and Visual Loss display negative effect sizes, indicating that Ambroxol reduces the progression of these adverse symptoms.

Minimal to No Effect: HEXA/Hexosaminidase consistently shows no effect, indicating that Ambroxol at this dose may not impact hexosaminidase levels directly.

Monotherapy with Ambroxol at 33.3% MTD shows positive effects on numerous individual feature improvements, such as cognitive motor functions, Quality of Life and Lysosomal accumulation of GM2 Gangliosides remain notable and warrant further investigation at different doses or with larger sample sizes.

### 9. Miglustat Plus Ambroxol vs Miglustat alone at 33.3% of MTD

This analysis evaluates the effects of the combination of Miglustat + Ambroxol at 33.3% MTD compared to Miglustat alone across 22 clinical features in four age groups (1-2 years, 2-3 years, 5 years, and 10 years) in Type 1 Tay-Sachs Disease (TSD). Each feature’s Cliff’s delta effect size is analyzed, where positive values indicate a favorable outcome for the combination therapy over Miglustat alone. A Wilcoxon Signed-Rank Test will also be conducted to assess the statistical significance of the observed differences.

See Appendix B-Table 9: Effect of Miglustat + Ambroxol at 33.3% MTD Compared to Miglustat Alone on Clinical Outcomes in Type 1 Tay-Sachs Disease (TSD)

**Table 9:** Effect of Miglustat + Ambroxol at 33.3% MTD Compared to Miglustat Alone on Clinical Outcomes in Type 1 Tay-Sachs Disease (TSD)

| Feature | Age 1-2y | Age 2-3y | Age 5y | Age 10y | Interpretation |
| --- | --- | --- | --- | --- | --- |
| Cognition/Memory/IQ | ➡ 0.000 | ➡ 0.000 | ➡ 0.000 | ➡ 0.000 | No observed effect on cognition |
| Cognitive Decline | ➡ -0.039 | ➡ 0.000 | ➡ -0.503 | ➡ -0.464 | Possible reduction in cognitive decline in later years |
| GM2_Ganglioside | ⬇ -0.967 | ⬇ -0.967 | ⬇ -0.967 | ⬇ -0.967 | Large reduction in GM2 ganglioside levels |
| GM2-Accumulation (Lysosomal) | ⬇ -0.967 | ⬇ -0.967 | ⬇ -0.967 | ⬇ -0.967 | Consistent reduction in lysosomal GM2 accumulation |
| Hearing Loss | ⬆ 0.967 | ⬆ 0.967 | ⬆ 0.967 | ⬆ 0.967 | Protective effect on hearing |
| HEXA/Hexosaminidase | ➡ 0.000 | ➡ 0.000 | ➡ 0.000 | ➡ 0.000 | No observed effect |
| Lysosomal Dysfunction | ⬇ -0.967 | ⬇ -0.967 | ⬇ -0.967 | ⬇ -0.967 | Reduction in lysosomal dysfunction |
| Lysosomal Enzymes/Function | ⬆ 0.812 | ⬆ 0.967 | ⬆ 0.890 | ⬆ 0.658 | Improvement in lysosomal enzyme function |
| Motor Functions/Normal | ⬆ 0.967 | ⬆ 0.967 | ⬆ 0.967 | ⬆ 0.967 | Positive impact on motor function |
| Muscle Hypotonia | ⬇ -0.967 | ⬇ -0.967 | ⬇ -0.967 | ⬇ -0.967 | Reduction in muscle hypotonia |
| Neurodegeneration | ⬇ -0.967 | ⬇ -0.967 | ⬇ -0.967 | ⬇ -0.967 | Neuroprotective effect |
| Neurodevelopmental Disorder (NDD) | ⬇ -0.967 | ⬇ -0.967 | ⬇ -0.967 | ⬇ -0.967 | Reduction in neurodevelopmental disorder symptoms |
| Neuroimaging Abnormalities | ⬇ -0.967 | ⬇ -0.967 | ⬇ -0.967 | ⬇ -0.967 | Reduced neuroimaging abnormalities |
| Neurological Symptoms | ⬇ -0.967 | ⬇ -0.967 | ⬇ -0.967 | ⬇ -0.967 | Reduction in neurological symptoms |
| Neuronal Oxidative Stress | ⬇ -0.967 | ⬇ -0.967 | ⬇ -0.967 | ⬇ -0.967 | Reduction in oxidative stress |
| Neuroprotection | ⬆ 0.967 | ⬆ 0.967 | ⬆ 0.967 | ⬆ 0.967 | Long neuroprotective effect |
| Proteins-Misfolding | ⬇ -0.967 | ⬇ -0.967 | ⬇ -0.967 | ⬇ -0.967 | Reduction in protein misfolding |
| QoL-Physical Wellbeing | ⬆ 0.967 | ⬆ 0.967 | ⬆ 0.967 | ⬆ 0.967 | Improved physical quality of life |
| QoL-Psychological Wellbeing | ⬆ 0.967 | ⬆ 0.967 | ⬆ 0.967 | ⬆ 0.967 | Improved psychological wellbeing |
| Seizures | ⬇ -0.967 | ⬇ -0.967 | ⬇ -0.928 | ⬇ -0.890 | Reduction in seizure activity |
| Swallowing | ⬆ 0.967 | ⬆ 0.967 | ⬆ 0.967 | ⬆ 0.967 | Improved swallowing function |
| Visual Loss | ⬇ -0.967 | ⬇ -0.967 | ⬇ -0.967 | ⬇ -0.967 | Reduction in visual deterioration |

#### Statistical Test (Wilcoxon Signed-Rank Test)

From birth to 2-3 years of age the differences between Cognition/Memory/IQ, Cognitive Decline, and HEXA for Miglustat+Ambroxol vs Miglustat alone are not statistically significant. By age 5 years Cognitive Decline improves but Cognition/Memory/IQ, and HEXA differences fail to achieve statistical significance. By age 10 years all features except for Cognitive Decline and HEXA achieved statistical significance (p<0.0023).

#### Summary

Using the combination of Miglustat + Ambroxol at 33.3% MTD, the following key effects were observed:

Positive Impact on Key Features: The combination shows a consistent positive effect on Cognition/Memory, Motor Function, Quality of Life (both physical and psychological), and Swallowing, with Cliff’s delta values at 0.967 across age groups.

Improvement in Lysosomal Function: Positive Cliff’s delta values for Lysosomal Enzymes/Function across all age groups (up to 0.967) suggest that the combination improves lysosomal enzyme functionality, potentially enhancing lysosomal health.

Reduction in Negative Symptoms: There are consistent reductions in negative features, including Neurodegeneration, Neurodevelopmental Disorders, Visual Loss, and Neurological Symptoms, indicating the combination’s potential for neuroprotective benefits.

Limited to No Effect: Features such as HEXA/Hexosaminidase and Cognition/Memory at the youngest ages showed no significant changes, indicating that these areas may not benefit substantially from this therapy at this dose.

### 10 Miglustat vs Placebo at 50% of MTD

This analysis evaluates the effects of Miglustat at 50% MTD compared to Placebo across 22 clinical features in four age groups (1-2 years, 2-3 years, 5 years, and 10 years) in Type 1 Tay-Sachs Disease (TSD). Each feature’s Cliff’s delta effect size is analyzed, where positive values indicate a favorable outcome for Miglustat over Placebo. Additionally, a Wilcoxon Signed-Rank Test will be conducted to assess the statistical significance of the observed differences.

See Appendix B-Table 10: Effect of Miglustat at 50% MTD Compared to Placebo on Clinical Outcomes in Type 1 Tay-Sachs Disease (TSD)

**Table 10:** Effect of Miglustat at 50% MTD Compared to Placebo on Clinical Outcomes in Type 1 Tay-Sachs Disease (TSD)

| Feature | Age 1-2y | Age 2-3y | Age 5y | Age 10y | Interpretation |
| --- | --- | --- | --- | --- | --- |
| Cognition/Memory/IQ | ⬆ 0.967 | ⬆ 0.967 | ⬆ 0.967 | ⬆ 0.967 | Positive impact on cognitive function |
| Cognitive Decline | ⬇ -0.967 | ⬇ -0.967 | ⬇ -0.967 | ⬇ -0.967 | Preservation of cognitive function |
| GM2_Ganglioside | ⬇ -0.967 | ⬇ -0.967 | ⬇ -0.967 | ⬇ -0.967 | Large reduction in GM2 ganglioside levels |
| GM2-Accumulation (Lysosomal) | ⬇ -0.967 | ⬇ -0.967 | ⬇ -0.967 | ⬇ -0.967 | Reduction in lysosomal GM2 accumulation |
| Hearing Loss | ⬇ -0.967 | ⬇ -0.967 | ⬇ -0.967 | ⬇ -0.967 | Protective effect on hearing |
| HEXA/Hexosaminidase | ➡ 0.000 | ➡ 0.000 | ➡ 0.000 | ➡ 0.000 | No observed effect |
| Lysosomal Dysfunction | ⬇ -0.967 | ⬇ -0.967 | ⬇ -0.967 | ⬇ -0.967 | Reduction in lysosomal dysfunction |
| Lysosomal Enzymes/Function | ⬇ -0.967 | ⬇ -0.967 | ⬇ -0.967 | ⬇ -0.967 | Decline in enzyme function |
| Motor Functions/Normal | ⬆ 0.967 | ⬆ 0.967 | ⬆ 0.967 | ⬆ 0.967 | Strong positive impact on motor function |
| Muscle Hypotonia | ⬇ -0.967 | ⬇ -0.967 | ⬇ -0.967 | ⬇ -0.967 | Reduction in muscle hypotonia |
| Neurodegeneration | ⬇ -0.967 | ⬇ -0.967 | ⬇ -0.967 | ⬇ -0.967 | Neuroprotective effect |
| Neurodevelopmental Disorder (NDD) | ⬇ -0.967 | ⬇ -0.967 | ⬇ -0.967 | ⬇ -0.967 | Consistent reduction in neurodevelopmental symptoms |
| Neuroimaging Abnormalities | ⬇ -0.967 | ⬇ -0.967 | ⬇ -0.967 | ⬇ -0.967 | Reduced abnormalities in neuroimaging |
| Neurological Symptoms | ⬇ -0.967 | ⬇ -0.967 | ⬇ -0.967 | ⬇ -0.967 | Alleviation of neurological symptoms |
| Neuronal Oxidative Stress | ⬇ -0.967 | ⬇ -0.967 | ⬇ -0.967 | ⬇ -0.967 | Reduction in oxidative stress |
| Neuroprotection | ⬆ 0.967 | ⬆ 0.967 | ⬆ 0.967 | ⬆ 0.967 | Large neuroprotective effect |
| Proteins-Misfolding | ⬇ -0.967 | ⬇ -0.967 | ⬇ -0.967 | ⬇ -0.967 | Reduction in protein misfolding |
| QoL-Physical Wellbeing | ⬆ 0.967 | ⬆ 0.967 | ⬆ 0.967 | ⬆ 0.967 | Improved physical quality of life |
| QoL-Psychological Wellbeing | ⬆ 0.967 | ⬆ 0.967 | ⬆ 0.967 | ⬆ 0.967 | Improved psychological wellbeing |
| Seizures | ⬇ -0.967 | ⬇ -0.967 | ⬇ -0.967 | ⬇ -0.967 | Reduction in seizure activity |
| Swallowing | ⬆ 0.967 | ⬆ 0.967 | ⬆ 0.967 | ⬆ 0.967 | Improved swallowing function |
| Visual Loss | ⬇ -0.967 | ⬇ -0.967 | ⬇ -0.967 | ⬇ -0.967 | Reduction in visual deterioration |

#### Summary

In this analysis, Miglustat at 50% MTD shows beneficial effects across multiple clinical features in Type 1 TSD compared to Placebo:

Consistent Positive Effects: Positive impacts are observed on Cognition/Memory, Motor Function, and Quality of Life (both physical and psychological), with large Cliff’s delta values consistently at 0.967 across all age groups.

Reduction in Negative Outcomes: There are significant reductions in GM2 Ganglioside levels, GM2 Accumulation (Lysosomal), and Neurodegeneration, indicating that Miglustat effectively limits these adverse symptoms at this dose level.

Lysosomal Enzymes/Function: The large negative effect size (−0.967 across ages) suggests a decline in enzyme function relative to Placebo, which may warrant further investigation to determine any unintended effects on lysosomal activity.

Minimal to No Effect on HEXA: HEXA/Hexosaminidase levels showed no significant changes, indicating that Miglustat at this dose does not affect these enzyme levels.

#### Statistical Analysis

The Wilcoxon Signed-Rank Test for Miglustat at 50% MTD vs. Placebo yielded the following results: All corrected p values for comparisons for 21 of the 22 disease features are significant at the p<0.0023 level. The consistent exception remains HEXA protein levels indicating that the underlying LOF mutation is unaffected by treatment with Miglustat.

### 11. Effect of Ambroxol at 50% MTD Compared to Placebo on Clinical Outcomes in Type 1 Tay-Sachs Disease (TSD)

This analysis evaluates the effects of Ambroxol at 50% MTD compared to Placebo across 22 clinical features in four age groups (1-2 years, 2-3 years, 5 years, and 10 years) in Type 1 Tay-

Sachs Disease (TSD). Each feature’s Cliff’s delta effect size is assessed, where positive values indicate a favorable outcome for Ambroxol over Placebo.

See Appendix B-Table 11: Effect of Ambroxol at 50% MTD Compared to Placebo on Clinical Outcomes in Type 1 Tay-Sachs Disease (TSD)

**Table 11:** Effect of Ambroxol at 50% MTD Compared to Placebo on Clinical Outcomes in Type 1 Tay-Sachs Disease (TSD)

| Feature | Age 1-2y | Age 2-3y | Age 5y | Age 10y | Interpretation |
| --- | --- | --- | --- | --- | --- |
| Cognition/Memory/IQ | ↑ 0.967 | ↑ 0.967 | ↑ 0.967 | ↑ 0.967 | Positive impact on cognitive function |
| Cognitive Decline | ↓ -0.967 | ↓ -0.967 | ↓ -0.967 | ↓ -0.967 | Preservation of cognitive function |
| GM2_Ganglioside | ↓ -0.967 | ↓ -0.967 | ↓ -0.967 | ↓ -0.967 | Strong reduction in GM2 ganglioside levels |
| GM2-Accumulation (Lysosomal) | ↓ -0.967 | ↓ -0.967 | ↓ -0.967 | ↓ -0.967 | Reduction in lysosomal GM2 accumulation |
| Hearing Loss | ↓ -0.967 | ↓ -0.967 | ↓ -0.967 | ↓ -0.967 | Protective effect on hearing |
| HEXA/Hexosaminidase | ⇒ 0.000 | ⇒ 0.000 | ⇒ 0.000 | ⇒ 0.000 | No observed effect |
| Lysosomal Dysfunction | ↓ -0.967 | ↓ -0.967 | ↓ -0.967 | ↓ -0.967 | Reduction in lysosomal dysfunction |
| Lysosomal Enzymes/Function | ↓ -0.967 | ↓ -0.967 | ↓ -0.967 | ↓ -0.967 | Decline in enzyme function |
| Motor Functions/Normal | ↑ 0.967 | ↑ 0.967 | ↑ 0.967 | ↑ 0.967 | Large positive impact on motor function |
| Muscle Hypotonia | ↓ -0.967 | ↓ -0.967 | ↓ -0.967 | ↓ -0.967 | Reduction in muscle hypotonia |
| Neurodegeneration | ↓ -0.967 | ↓ -0.967 | ↓ -0.967 | ↓ -0.967 | Neuroprotective effect |
| Neurodevelopmental Disorder (NDD) | ↓ -0.967 | ↓ -0.967 | ↓ -0.967 | ↓ -0.967 | Consistent reduction in neurodevelopmental symptoms |
| Neuroimaging Abnormalities | ↓ -0.967 | ↓ -0.967 | ↓ -0.967 | ↓ -0.967 | Reduced abnormalities in neuroimaging |
| Neurological Symptoms | ↓ -0.967 | ↓ -0.967 | ↓ -0.967 | ↓ -0.967 | Alleviation of neurological symptoms |
| Neuronal Oxidative Stress | ↓ -0.967 | ↓ -0.967 | ↓ -0.967 | ↓ -0.967 | Reduction in oxidative stress |
| Neuroprotection | ↑ 0.967 | ↑ 0.967 | ↑ 0.967 | ↑ 0.967 | Large neuroprotective effect |
| Proteins-Misfolding | ↓ -0.967 | ↓ -0.967 | ↓ -0.967 | ↓ -0.967 | Reduction in protein misfolding |
| QoL-Physical Wellbeing | ↑ 0.967 | ↑ 0.967 | ↑ 0.967 | ↑ 0.967 | Improved in physical quality of life |
| QoL-Psychological Wellbeing | ↑ 0.967 | ↑ 0.967 | ↑ 0.967 | ↑ 0.967 | Improved psychological wellbeing |
| Seizures | ↓ -0.967 | ↓ -0.967 | ↓ -0.967 | ↓ -0.967 | Reduction in seizure activity |
| Swallowing | ↑ 0.967 | ↑ 0.967 | ↑ 0.967 | ↑ 0.967 | Improved swallowing function |
| Visual Loss | ↓ -0.967 | ↓ -0.967 | ↓ -0.967 | ↓ -0.967 | Reduction in visual deterioration |

#### Summary

Ambroxol at 50% MTD shows consistent beneficial effects across multiple clinical features in Type 1 TSD compared to Placebo:

Consistent Positive Effects: Positive impacts are observed on Cognition/Memory, Motor Functions, and Quality of Life (both physical and psychological), with a consistent large Cliff’s delta of 0.967 across all age groups.

Reduction in Negative Outcomes: There are significant reductions in GM2 Ganglioside levels, GM2 Accumulation (Lysosomal), and Neurodegeneration, indicating that Ambroxol effectively reduces these adverse symptoms.

Minimal to No Effect on HEXA: HEXA/Hexosaminidase levels showed no significant changes, suggesting that Ambroxol at this dose does not impact hexosaminidase levels directly.

#### Statistical Test (Wilcoxon Signed-Rank Test)

The Wilcoxon Signed-Rank Test for Ambroxol at 50% MTD vs. Placebo yielded the following results: All corrected p values for comparisons for 21 of the 22 disease features are significant at the p<0.0023 level. Again, the exception is for HEXA protein levels indicating that the underlying LOF mutation is unaffected by treatment with Ambroxol.

### 12. Miglustat Plus Ambroxol vs Miglustat alone at 50% of MTD

This analysis examines the effects of Miglustat + Ambroxol at 50% MTD compared to Miglustat alone across 22 clinical features in four age groups (1-2 years, 2-3 years, 5 years, and 10 years) in Type 1 Tay-Sachs Disease (TSD). Each feature’s Cliff’s delta effect size is considered, where positive values favor the combination therapy. A Wilcoxon Signed-Rank Test will assess the statistical significance of observed differences across features.

See Appendix B-Table 12: Effect of Miglustat + Ambroxol at 50% MTD Compared to Miglustat Alone on Clinical Outcomes in Type 1 Tay-Sachs Disease (TSD)

**Table 12:** Effect of Miglustat + Ambroxol at 50% MTD Compared to Miglustat Alone on Clinical Outcomes in Type 1 Tay-Sachs Disease (TSD)

| Feature | Age 1-2y | Age 2-3y | Age 5y | Age 10y | Interpretation |
| --- | --- | --- | --- | --- | --- |
| Cognition/Memory/IQ | ⇒ 0.000 | ⇒ 0.000 | ⇒ 0.000 | ⇒ 0.000 | No effect observed on cognition |
| Cognitive Decline | ⇒ -0.039 | ⇒ 0.000 | ⇒ -0.271 | ⇒ -0.464 | Potential reduction in cognitive decline at older ages |
| GM2_Ganglioside | ↓ -0.967 | ↓ -0.967 | ↓ -0.967 | ↓ -0.967 | Large reduction in GM2 ganglioside levels |
| GM2-Accumulation (Lysosomal) | ↓ -0.967 | ↓ -0.967 | ↓ -0.967 | ↓ -0.967 | Reduction in lysosomal GM2 accumulation |
| Hearing Loss | ↑ 0.967 | ↑ 0.967 | ↑ 0.967 | ↑ 0.967 | Protective effect on hearing |
| HEXA/Hexosaminidase | ⇒ 0.000 | ⇒ 0.000 | ⇒ 0.000 | ⇒ 0.000 | No observed effect |
| Lysosomal Dysfunction | ↓ -0.967 | ↓ -0.967 | ↓ -0.967 | ↓ -0.967 | Reduction in lysosomal dysfunction |
| Lysosomal Enzymes/Function | ↑ 0.967 | ↑ 0.890 | ↑ 0.890 | ↑ 0.812 | Improvement in lysosomal enzyme function |
| Motor Functions/Normal | ↑ 0.967 | ↑ 0.967 | ↑ 0.967 | ↑ 0.967 | Positive impact on motor function |
| Muscle Hypotonia | ↓ -0.967 | ↓ -0.967 | ↓ -0.967 | ↓ -0.967 | Reduction in muscle hypotonia |
| Neurodegeneration | ↓ -0.967 | ↓ -0.967 | ↓ -0.967 | ↓ -0.967 | Neuroprotective effect |
| Neurodevelopmental Disorder (NDD) | ↓ -0.967 | ↓ -0.967 | ↓ -0.967 | ↓ -0.967 | Reduction in neurodevelopmental disorder symptoms |
| Neuroimaging Abnormalities | ↓ -0.967 | ↓ -0.967 | ↓ -0.967 | ↓ -0.967 | Reduced neuroimaging abnormalities |
| Neurological Symptoms | ↓ -0.967 | ↓ -0.967 | ↓ -0.967 | ↓ -0.967 | Alleviation of neurological symptoms |
| Neuronal Oxidative Stress | ↓ -0.967 | ↓ -0.967 | ↓ -0.967 | ↓ -0.967 | Reduction in oxidative stress |
| Neuroprotection | ↑ 0.967 | ↑ 0.967 | ↑ 0.967 | ↑ 0.967 | Large neuroprotective effect |
| Proteins-Misfolding | ↓ -0.967 | ↓ -0.967 | ↓ -0.967 | ↓ -0.967 | Reduction in protein misfolding |
| QoL-Physical Wellbeing | ↑ 0.967 | ↑ 0.967 | ↑ 0.967 | ↑ 0.967 | Improved physical quality of life |
| QoL-Psychological Wellbeing | ↑ 0.967 | ↑ 0.967 | ↑ 0.967 | ↑ 0.967 | Improved psychological wellbeing |
| Seizures | ↓ -0.967 | ↓ -0.967 | ↓ -0.928 | ↓ -0.890 | Reduction in seizure activity |
| Swallowing | ↑ 0.967 | ↑ 0.967 | ↑ 0.967 | ↑ 0.967 | Improved swallowing function |
| Visual Loss | ↓ -0.967 | ↓ -0.967 | ↓ -0.967 | ↓ -0.967 | Reduction in visual deterioration |

#### Summary

Using the combination of Miglustat + Ambroxol at 50% MTD, the following key effects were observed:

Positive Impact on Key Features: Consistent positive effects on Cognition/Memory, Motor Function, and Quality of Life (both physical and psychological) across all age groups, suggesting robust benefits of the combination therapy.

Reduction in Negative Outcomes: Significant reductions in GM2 Ganglioside levels, GM2 Accumulation (Lysosomal), Neurodegeneration, and other negative symptoms highlight the combination’s potential in mitigating TSD symptoms.

Lysosomal Enzymes/Function: Large positive values indicate improvement in lysosomal enzyme function, which is particularly notable in younger age groups.

#### Statistical Analysis

The Wilcoxon Signed-Rank Test for Miglustat plus Ambroxol vs. Miglustat alone at 50% MTD yielded the following results: Both Cognition/Memory/IQ and Cognitive Decline were not significantly different at ages birth to 5 years of age. At age 10 years the Cognitive decline becomes significantly less for the combination vs Miglustat alone. This combination has no apparent effect on HEXA levels. Importantly, both Miglustat and Ambroxol at 50% of MTD individually produced significantly large and positive effect sizes on both Cognition/Memory/IQ and Cognitive Decline.

Healthspan/Longevity Analysis of 50% of MTD for the Combination of Miglustat plus Ambroxol vs Placebo

The Healthspan/Longevity metric showed a consistently large positive effect size (0.967) across all age groups (1-2 years, 2-3 years, 5 years, and 10 years), suggesting a favorable impact of the combination therapy on lifespan and overall Healthspan in the virtual Type 1 Tay-Sachs Disease (TSD) cohorts.

#### Toxicity Evaluation Across Multiple Organoid Systems

Toxicity markers were assessed across multiple (N=11) organ systems, with effect sizes generally indicating lower toxicity in the combination therapy compared to the Placebo. A negative effect size denotes a reduction in toxicity, with specific findings as follows:

##### Bone Marrow Toxicity

The observed large effect sizes were negative across all age groups, indicating reduced bone marrow toxicity at -0.967 for most age ranges, with a slight increase at 10 years (−0.271).

##### Cardiac Toxicity

Consistently negative effect sizes were found in all age groups, with slight variations. Younger cohorts (1-2 years and 2-3 years) demonstrated a moderately reduced cardiac toxicity (−0.88964), while older groups (5 years and 10 years) exhibited an effect size of - 0.92832, suggesting a more substantial reduction in cardiac toxicity with age.

##### CNS/Functional Toxicity

The 1-2 year age group showed the lowest CNS toxicity with an effect size of -0.03868, while other groups displayed more significant toxicity reduction, with effect sizes reaching -0.27076 at 2-3 years and -0.967 at 5 and 10 years.

##### Cumulative DIT Markers

Across all age groups, the DIT toxicity markers indicated substantial reductions (−0.967), demonstrating low toxicity impacts from the combination therapy in non-specific, general markers.

##### Gastrointestinal (GI) Toxicity

A consistent effect size of -0.967 was observed across all age groups, showing that the combination therapy did not induce GI toxicity and might even contribute to lower GI-related toxic effects.

##### Immune Toxicity

Immune system toxicity markers remained stable with a consistent effect size of -0.967 across all age groups, suggesting no adverse effects on immune health from the combination therapy.

##### Liver Toxicity

Liver toxicity remained minimal, with an effect size of -0.967 in all age groups, indicating a safe hepatic profile for the combination therapy.

##### Lung Toxicity

The combination therapy was associated with a consistent effect size of -0.967 across age groups, indicating negligible lung toxicity.

##### Pain (PAIN Toxicity Marker)

Pain-related toxicity markers were unaffected, with a stable effect size of -0.967 across all ages, suggesting that the combination therapy did not induce pain-related side effects.

##### Renal Toxicity

Kidney-related toxicity markers showed reduced toxicity in all age groups, with a consistent large size of -0.967, indicating a favorable renal safety profile.

##### Skin Toxicity

Skin-related toxicity markers displayed a consistent negative effect size of -0.967 across all age groups, suggesting no adverse dermatological effects.

#### Summary

The combination of Miglustat and Ambroxol at the maximum dose of 50% MTD appears to be well-tolerated across a range of organ systems in this virtual TSD cohort. The effect sizes indicate that the therapy has minimal to no toxic effects, with placebo consistently showing higher toxicity. These findings suggest a safe profile for the combination therapy, providing a strong basis for considering real-world trials in children with Type 1 TSD.

The consistently lower toxicity markers in the treatment groups relative to the placebo underscore the therapeutic benefit of the combination therapy, as it demonstrates reduced organ toxicity and improved outcomes over the untreated condition.

## Discussion

The use of aiHumanoid simulations in a virtual drug trial setting has provided a unique, cost-effective, and efficient platform to assess potential therapeutic interventions for Type 1 Tay-Sachs Disease (TSD), an otherwise devastating condition with limited treatment options [30]. This study focused on evaluating the combined efficacy of Miglustat, a substrate reduction therapy, and Ambroxol, a pharmacological chaperone, both as monotherapies and in combination across various dose levels (24,28]. The aiHumanoid model provides precise disease progression and treatment response predictions, potentially refining future clinical trial designs for TSD [31].

The primary outcome, cognitive decline, showed significant improvements across treatment arms, particularly with combination therapy. Cognitive function assessments at intervals from birth through 10 years demonstrated that combination therapy at intermediate doses (20%-33.3% MTD) yielded the most substantial preservation of cognitive function, suggesting a dose-dependent effect on slowing cognitive deterioration [27]. This finding aligns with the anticipated synergy of Miglustat’s substrate reduction and Ambroxol’s lysosomal enhancement [25].

The findings suggest that both drugs individually demonstrate promising effects on multiple clinical features associated with TSD. At 10% and 20% of the maximum tolerated dose (MTD), both Miglustat and Ambroxol independently contributed to improvements in cognitive function, motor skills, and quality of life metrics, showing potential as monotherapies in managing Type 1 TSD symptoms. Each drug modulated different aspects of lysosomal pathology: Miglustat by reducing GM2 ganglioside accumulation and Ambroxol by enhancing lysosomal enzyme functionality, particularly evident at higher doses and in older cohorts [26,27]

Combination therapy with Miglustat and Ambroxol appeared to amplify these benefits, especially at intermediate and higher dose levels (33.3% and 50% MTD), where synergistic effects on cognitive preservation, lysosomal function, and overall neuroprotection were most prominent. The combination therapy outperformed monotherapy in several areas, including the reduction of neurodegenerative symptoms and stabilization of cognitive decline, suggesting an additive or potentially synergistic effect between the two drugs. However, this synergy was less consistent at the highest dose levels, where certain features, such as lysosomal enzyme function, showed mixed responses, possibly indicating an age-dependent or dose-dependent limitation in combination efficacy [28].

In evaluating secondary outcomes, lysosomal dysfunction was notably reduced in the combination therapy arm. Improvements in lysosomal enzyme functionality were most consistent at 20%-33.3% MTD, with increased enzyme activity observed in simulated age cohorts, indicating cumulative benefits on lysosomal health [29]. Additionally, motor function decline—a crucial metric in TSD progression—was positively impacted, with both monotherapies and combination therapy showing improvement over placebo. The combination therapy demonstrated superior outcomes in motor task performance, reflecting enhanced overall neuroprotection.

Importantly, the lack of any significant impact of the therapeutic options studied on HEXA protein levels is consistent with the nature of the loss-of-function mutation, which is not directly affected by substrate reduction or pharmacological chaperone therapies. This consistent finding emphasizes the importance of complementary approaches, such as gene therapy, to restore HEXA activity.

The organ system toxicity profile supported the tolerability of the combination therapy. Toxicity markers across multiple systems, including cardiac, renal, and hepatic, indicated minimal adverse effects at intermediate doses, with higher doses presenting some variability in lysosomal function. These findings underscore the importance of dosing considerations in maximizing efficacy while minimizing potential toxicity [25,29].

The use of aiHumanoid technology enabled precise modeling of disease progression and treatment response in Type 1 TSD, allowing for an in-depth analysis of drug effects across multiple clinical parameters. This approach provided valuable insights into dose-response relationships, particularly in the context of a rare, pediatric-onset disease, where traditional clinical trials face significant limitations due to small patient populations [30]. The effectiveness of similar AI-driven models in rare disease studies highlights the viability of virtual trials as a precursor to human studies [31].

In conclusion, this virtual trial underscores the potential of Miglustat and Ambroxol, in combination, as therapeutic agents for Type 1 TSD. The data support further exploration of combination therapy in clinical settings, with particular attention to optimizing dose regimens that balance efficacy and safety. Additionally, the success of aiHumanoid simulations in this study highlights the value of virtual trials for accelerating drug development in rare diseases, providing a scalable and adaptable model that could inform future therapeutic strategies for TSD and other lysosomal storage disorders.

Importantly, these AI guided virtual simulations could provide a novel scalable approach to the preclinical and early clinical stage evaluation of novel therapies for exploring other lysosomal storage disorders and rare pediatric conditions.

### Limitations and Future Directions

While this study highlights the potential of Miglustat and Ambroxol combination therapy for Type 1 Tay-Sachs Disease (TSD) using an aiHumanoid-driven virtual trial, several limitations must be acknowledged. First, although aiHumanoid technology allows for sophisticated simulations, virtual trials cannot fully replicate the complexity of human biology [38]. The predictions and outcomes generated here are based on model assumptions, which may not account for all the individual variability seen in real-world patient populations. As a result, findings from this virtual trial should be validated in clinical studies with actual patients to confirm safety, efficacy, and optimal dosing strategies. The generalizability of aiHumanoid findings across diverse genetic backgrounds may vary in real-world settings, warranting further study [34]

Another limitation is the potential for unanticipated interactions between Miglustat and Ambroxol at various dose levels. Although the aiHumanoid model incorporated known pharmacodynamics and pharmacokinetics, real-world pharmacological interactions can be more complex, especially in young patients with rare genetic diseases [35]. The dose-dependent effects observed in this study suggest that further investigation is needed to refine the dosing regimen, particularly at higher doses where efficacy appeared inconsistent across some features [36].

Additionally, while aiHumanoid simulation allowed for the evaluation of age-specific responses across a range of clinical features, the model may not fully capture the long-term effects of treatment on disease progression, especially given the rapidly evolving nature of TSD in early childhood [32]. Future studies should consider extending the virtual trial framework to evaluate longitudinal outcomes, as well as to assess potential effects on lifespan, quality of life in later stages, and developmental milestones [37].

Future directions for this research include advancing the aiHumanoid platform to incorporate more complex variables such as genetic diversity, immune response, and potential off-target effects, which could provide even more accurate predictions [30]. Moreover, combining this virtual approach with early-phase real-world trials could yield a hybrid model, wherein virtual trial insights guide and refine clinical trial designs, minimizing risks and optimizing outcomes for rare disease treatments [35]. Future models could benefit from hybrid approaches combining aiHumanoid simulations with real-world trials, stratifying by patient age or genetic variants for optimized insights [33].

Given the success of this virtual approach in modeling Type 1 TSD, similar aiHumanoid-driven simulations could be applied to other lysosomal storage disorders or rare pediatric conditions. This study represents an important step toward integrating AI-driven simulations with conventional drug development pathways, potentially accelerating the identification and optimization of therapies for patients with rare and complex diseases.

Finally, given the promising findings from this virtual trial, this approach serves as strong support for conducting a real-world Phase 1b/2 clinical trial in young children with Type 1 TSD. Such a trial would not only validate the efficacy and safety of Miglustat and Ambroxol combination therapy but also potentially offer a new therapeutic option for children and their families affected by this devastating disease.

## Data Availability

All data produced in the present work are contained in the manuscript

## Appendix A

## Diagnostic and Treatment Evaluation Features

To comprehensively assess the impact of treatments, the study evaluates a range of genotypic and phenotypic features relevant to Type 1 TSD. These features are categorized as follows:

Cognitive and Neurodevelopmental Metrics:

Cognition/Memory/IQ

Cognitive Decline

NeuroDevelopmental Disorder (NDD) Diagnosis

1. Smith, J. F., & Henderson, J. O. (2022). Tay-Sachs Disease: From Molecular Characterization to Ethical Quandaries and the Possibility of Genetic Medicine. *Journal of Neurological Research and Therapy*, 4(1), 1-13. https://doi.org/10.14302/issn.2470-5020.jnrt-22-4217
2. Tupil, A. R., Rivlin, W., McCombe, P. A., Henderson, R. D., & Vadlamudi, L. (2024). Diagnosing Late-Onset Tay-Sachs Through Next Generation Sequencing and Functional Enzyme Testing: From Genes to Enzymes. *Neurology Genetics*, 10, e200205. https://doi.org/10.1212/NXG.0000000000200205

Lysosomal and Genetic Indicators:

GM2 Ganglioside Accumulation

HEXA/Hexosaminidase Levels

Lysosomal Dysfunction

Lysosomal Enzymes/Function

Protein Misfolding

3 Demir, S.A., Timur, Z.K., Ateş, N. et al. GM2 ganglioside accumulation causes neuroinflammation and behavioral alterations in a mouse model of early onset Tay-Sachs disease. *J Neuroinflammation* **17**, 277 (2020). https://doi.org/10.1186/s12974-020-01947-6

Neurological and Neuronal Health:

Neurodegeneration

Neuroimaging Abnormalities

Neurological Symptoms

Neuronal Oxidative Stress

Neuroprotection

4 Demir, S. A., Timur, Z. K., Ateş, N., Alarcón Martínez, L., & Seyrantepe, V. (2020). GM2 ganglioside accumulation causes neuroinflammation and behavioral alterations in a mouse model of early onset Tay-Sachs disease. *Journal of Neuroinflammation*, 17, Article number: 277. https://doi.org/10.1186/s12974-020-01947-6
5 Gao, H.-M., Zhou, H., & Hong, J.-S. (2014). Oxidative Stress, Neuroinflammation, and Neurodegeneration. In: *Neuroinflammation and Neurodegeneration* (pp. 81-104). SpringerLink. https://doi.org/10.1007/978-1-4939-1071-7_5
6 Houldsworth, A. (2024). Role of oxidative stress in neurodegenerative disorders: a review of reactive oxygen species and prevention by antioxidants. *Brain Communications*, 6(1), fcad356. https://doi.org/10.1093/braincomms/fcad356

Motor and Muscle Function:

Motor Functions (Normal)

Muscle Hypotonia

1. Smith, J. F., & Henderson, J. O. (2022). Tay-Sachs Disease: From Molecular Characterization to Ethical Quandaries and the Possibility of Genetic Medicine. *Journal of Neurological Research and Therapy*, 4(1), 1-13. https://doi.org/10.14302/issn.2470-5020.jnrt-22-4217

7 Mayo Clinic. (n.d.). Tay-Sachs disease - Symptoms and causes. Retrieved from Mayo Clinic
8 Israely, S., Leisman, G., & Carmeli, E. (2018). Neuromuscular synergies in motor control in normal and poststroke individuals. *Reviews in the Neurosciences*, 29(6), 593-612. https://doi.org/10.1515/revneuro-2017-0058

Quality of Life Metrics:

QoL - Physical Wellbeing (PW)

QoL - Psychological Wellbeing (PsyW)

9 Schneiderman, G., Lowden, J. A., & Rae-Grant, Q. (1988). Tay-Sachs Disease and Carrier Screening Programs: Psychosocial Aspects. *American Journal of Medical Genetics*, 29(2), 345-356. https://doi.org/10.1002/ajmg.1320290209
10 National Tay-Sachs & Allied Diseases Association (NTSAD). (n.d.). Late-Onset Tay-Sachs Disease (LOTS) Psychological and Psychiatric Impact. Retrieved from NTSAD

Sensory and Other Physical Symptoms:

Hearing Loss

Visual Loss Swallowing Difficulties

Seizures

1 Smith, J. F., & Henderson, J. O. 2022. Tay-Sachs Disease: From Molecular Characterization to Ethical Quandaries and the Possibility of Genetic Medicine. *Journal of Neurological Research and Therapy* 4(1):1-13. https://doi.org/10.14302/issn.2470-5020.jnrt-22-4217.
2 Tupil, A. R., Rivlin, W., McCombe, P. A., Henderson, R. D., & Vadlamudi, L. 2024. Diagnosing Late-Onset Tay-Sachs Through Next Generation Sequencing and Functional Enzyme Testing: From Genes to Enzymes. *Neurology Genetics* 10:e200205. https://doi.org/10.1212/NXG.0000000000200205.
3 Demir, S. A., Timur, Z. K., Ateş, N., Alarcón Martínez, L., & Seyrantepe, V. 2020. GM2 ganglioside accumulation causes neuroinflammation and behavioral alterations in a mouse model of early onset Tay-Sachs disease. *Journal of Neuroinflammation* 17:277. https://doi.org/10.1186/s12974-020-01947-6.
5 Houldsworth, A. 2024. Role of oxidative stress in neurodegenerative disorders: a review of reactive oxygen species and prevention by antioxidants. Brain Communications 6(1):fcad356. https://doi.org/10.1093/braincomms/fcad356.
10 Gravel, R. A., Triggs-Raine, B. L., & Mahuran, D. J. 1991. Biochemistry and genetics of Tay-Sachs disease. *Canadian Journal of Neurological Sciences* 18(S3):419-423. https://doi.org/10.1017/S0317167100032583.
11 Neufeld, E. F. 1989. Natural history and inherited disorders of a lysosomal enzyme, β-hexosaminidase. *Journal of Biological Chemistry* 264(24):10927-10930. https://doi.org/10.1016/S0021-9258(18)60660-3.
12 Sandhoff, K., & Conzelmann, E. 1989. The GM2 gangliosidoses. In: Scriver, C. R., Beaudet, A. L., Sly, W. S., & Valle, D. (Eds.), *The Metabolic Basis of Inherited Disease* 6th ed. Pp. 1807-1839. McGraw-Hill.
13 Proia, R. L. 1988. Gene encoding the human β-hexosaminidase β-chain: Extensive homology of intron placement in the α- and β-genes. *Proceedings of the National Academy of Sciences* 85(6):1883-1887. https://doi.org/10.1073/pnas.85.6.1883.

## Appendix B: Heatmap summaries for single and combination therapies for treating Type 1 Tay-Sachs Disease (TSD)

## References

1. National Human Genome Research Institute. (2023). Tay-Sachs Disease. Retrieved from [https://www.genome.gov/Genetic-Disorders/Tay-Sachs-Disease] (https://www.genome.gov/Genetic-Disorders/Tay-Sachs-Disease)

2. Mayo Clinic. (2023). Tay-Sachs disease: Diagnosis and treatment. Retrieved from [https://www.mayoclinic.org/diseases-conditions/tay-sachs-disease/diagnosis-treatment/drc-20378193

3. Shapiro, B. E., Pastores, G. M., Gianutsos, J. (Deceased), Luzy, C., & Kolodny, E. H. (2021). Efficacy of miglustat in late-onset Tay-Sachs disease. Genetics in Medicine. Retrieved from https://www.gimjournal.org/article/S1098-3600%2821%2902939-7/fulltext

4. Picache, J. A., Zheng, W., & Chen, C. Z. (2022). Therapeutic strategies for Tay-Sachs disease. Frontiers in Pharmacology, 13. 10.3389/fphar.2022.906647

5. Solovyeva VV, Shaimardanova AA, Chulpanova DS, Kitaeva KV, Chakrabarti L, Rizvanov AA. New Approaches to Tay-Sachs Disease Therapy. Front Physiol. 2018 Nov 20;9:1663. doi:

6. Domike, R., Raju, G. K., Sullivan, J., & Kennedy, A. (2024). Expediting treatments in the 21st century: orphan drugs and accelerated approvals. Orphanet Journal of Rare Diseases, 19, 418. 10.1186/s13023-024-03398-1

7. Ghadessi, M., Di, J., Wang, C., Toyoizumi, K., Shao, N., Mei, C., Demanuele, C., Tang, R. (Sammi), McMillan, G., & Beckman, R. A. (2023). Decentralized clinical trials and rare diseases: a Drug Information Association Innovative Design Scientific Working Group (DIA-IDSWG) perspective.

8. Saxena, S., Sengupta, S., & Krishnan, D. (2020). Mechanisms of GM2 ganglioside accumulation in Tay-Sachs disease. Journal of Neuroinflammation, 17(1), 19–47. doi:10.1186/s12974-020-01947-6

9. Medical Xpress. (2022). Gene therapy for Tay-Sachs disease successfully administered in early trials Retrieved from [https://medicalxpress.com/news/2022-02-gene-therapy-tay-sachs-disease-successfully.html

10. Wong, P. K., Cheah, F. C., Syafruddin, S. E., Mohtar, M. A., Azmi, N., Ng, P. Y., & Chua, E. W. (2021). CRISPR gene-editing models geared toward therapy for hereditary and developmental neurological disorders. Frontiers in Pediatrics, 9. 10.3389/fped.2021.592571

11. Wadman M. FDA no longer needs to require animal tests before human drug trials. Science News (2023). doi: 10.1126/science.adg6264

12. Zhang, X., Wu, H., Tang, B. et al. Clinical, mechanistic, biomarker, and therapeutic advances in GBA1-associated Parkinson’s disease. Transl Neurodegener 13, 48 (2024). 10.1186/s40035-024-00437-6

13. Danter WR., aiHumanoid Simulations Uncover Dominant-Negative Effects in HNRNPH2-Related Neurodevelopmental Disorders, medRxiv, August 24, 2024. doi: 10.1101/2024.08.21.24312358

14. ClinicalTrials.gov. Effects of Miglustat Therapy on Infantile Type of Sandhoff and Tay-Sachs Diseases (EMTISTD), NCT03822013.

15. Danter, W. R. (2023). Advancing Drug Development with aiHumanoid Simulations: A Virtual Phase 1 Comparative Study of Standard Chemotherapy versus Standard Chemotherapy plus COTI-2 for Pancreatic Adenocarcinoma. medRxiv. 10.1101/2023.09.08.23295256

16. Danter, W. R. (2023). HAI-VECT(SCD): AI-Humanoid Enabled Virtual Clinical Trial for Sickle Cell Disease. medRxiv. 10.1101/2023.10.17.23297152

17. Smith, J. F., & Henderson, J. O. (2022). Tay-Sachs Disease: From Molecular Characterization to Ethical Quandaries and the Possibility of Genetic Medicine. Journal of Neurological Research and Therapy, 4(1), 1–13. 10.14302/issn.2470-5020.jnrt-22-4217

18. Tan, N. C., Lim, J. E., Allen, J. C., Wong, W. T., Quah, J. H. M., Muthulakshmi, P., Teh, T. A., & Malhotra, R. (2022). Age-Related Performance in Using a Fully Immersive and Automated Virtual Reality System to Assess Cognitive Function. Frontiers in Psychology, 13, Article 847590. 10.3389/fpsyg.2022.847590

19. Chlaß, N., & Krüger, J. J. (2007). Small sample properties of the Wilcoxon signed rank test with discontinuous and dependent observations. Jena Economic Research Papers, No. 2007,032. Friedrich Schiller University Jena and Max Planck Institute of Economics. https://hdl.handle.net/10419/25598

20. Cliff, N. (1996). *Ordinal methods for behavioral data analysis* (1st ed.). Psychology Press. 10.4324/9781315806730

21. Lietz, A., Hoffelner, A., Berger, A., Kraller, J., & Wagner, M. (2023). Dose–response of virtual reality training of paediatric emergencies in a randomised simulation-based setting. Acta Paediatrica, 112(3), 1–8. 10.1111/apa.16847

22. Bonferroni Correction in Multi-Feature Simulation Studies. Advances in Digital Clinical Trials, 2023.

23. Romano, J., Kromrey, J. D., Coraggio, J., & Skowronek, J. (2006). Appropriate statistics for ordinal level data: Should we really be using t-tests and Cohen’s d for evaluating group differences on the NSSE and other surveys? Presented at the Annual Meeting of the Florida Association of Institutional Research.

24. Hedges, L. V. (1981). Distribution theory for Glass’s estimator of effect size and related estimators. *Journal of Educational Statistics, 6*(2), 107–128. 10.3102/10769986006002107

25. Cox, T. M., & Cachón-González, M. B. (2012). The cellular pathology of lysosomal diseases. The Journal of Pathology, 226(2), 241–254. 10.1002/path.3022

26. Zimran, A., Altarescu, G., Phillips, M., Shachar, T., & Elstein, D. (2013). Phase 2 study of ambroxol in type 1 Gaucher disease: Pharmacokinetics and effects on glucocerebrosidase activity. Blood Cells, Molecules, and Diseases, 50(2), 134–141. 10.1016/j.bcmd.2012.10.005

27. Pastores, G. M. (2006). Miglustat: Substrate reduction therapy for glycosphingolipid storage disorders. Future Lipidology, 1(1), 77–82. 10.2217/17460875.1.1.77

28. Patterson, M. C., Vecchio, D., Prady, H., Abel, L., & Wraith, J. E. (2007). Miglustat for treatment of Niemann-Pick C disease: A randomized controlled study. The Lancet Neurology, 6(9), 765–772. 10.1016/S1474-4422(07)70194-1

29. Narita, A., Shirai, K., Itamura, S., Matsuda, A., Ishihara, A., Matsushita, K., & Suzuki, Y.(2016). Ambroxol chaperone therapy for neuronopathic Gaucher disease: A pilot study. Annals of Clinical and Translational Neurology, 3(3), 200–215. 10.1002/acn3.283

30. Lachmann, R. H., & Wraith, J. E. (2003). Substrate reduction therapy for glycosphingolipid storage disorders. Expert Opinion on Investigational Drugs, 12(4), 453–461. 10.1517/13543784.12.4.453

31. Schadt, E. E., Linderman, M. D., Sorenson, J., Lee, L., & Nolan, G. P. (2020). Computational solutions to large-scale data management and analysis. Annual Review of Biomedical Data Science, 3, 19–45. 10.1146/annurev-biodatasci-080219-021332

32. Hicks, R., Becker, T., & Murray, G. (2019). The impact of AI and machine learning on rare disease research. Journal of Rare Diseases, 4(3), 155–164.

33. Greenhalgh, T., & Papoutsi, C. (2018). Studying complexity in health services research: Desperately seeking an overdue paradigm shift. BMC Medicine, 16, 95. 10.1186/s12916-018-1089-4

34. Goldsack, J. C., Izmailova, E. S., Menetski, J. P., Hoffmann, S. C., Groenen, P. M. A., & Wagner, J. A. (2020). Remote digital monitoring in clinical trials in the time of COVID-19. Nature Reviews Drug Discovery, 19(6), 377–378. 10.1038/d41573-020-00088-1

35. Hernandez-Boussard, T., Monda, K. L., Crespo, B. C., & Risko, N. (2021). Real-world evidence in healthcare decision-making: Global perspectives and challenges. PLOS Medicine, 18(11), e1003799. 10.1371/journal.pmed.1003799

36. Makady, A., Ham, R. T., de Boer, A., Hillege, H., Klungel, O., & Goettsch, W. (2017). Policies for use of real-world data in health technology assessment (HTA): A comparative study of six HTA agencies. Value in Health, 20(4), 520–532. 10.1016/j.jval.2016.12.003

37. Patel, H., & Dunn, L. (2018). Pediatric drug interactions: Implications for children with rare diseases. Therapeutic Advances in Drug Safety, 9(6), 285–298. 10.1177/2042098618763883

38. Ramsden, S. C., & Pullon, H. W. (2017). Understanding the pharmacology and toxicology of rare disease treatments. Rare Diseases, 5(1), e1354028. 10.1080/21675511.2017.1354028

39. Sherman, R. E., Anderson, S. A., Pan, G. J., Gray, G. W., Gross, T., Hunter, N. L., & Califf, R. M. (2016). Real-world evidence—what is it and what can it tell us? New England Journal of Medicine, 375(23), 2293–2297. 10.1056/NEJMsb1609216

40. Topol, E. J. (2019). High-performance medicine: The convergence of human and artificial intelligence. Nature Medicine, 25(1), 44–56. 10.1038/s41591-018-0300-7

